# Mapping total microbial communities and waterborne pathogens in household drinking water in China by citizen science and metabarcoding

**DOI:** 10.1101/2023.10.16.23297104

**Authors:** Xinyi Wen, Chutong Fang, Lihan Huang, Jiazheng Miao, Yajuan Lin

## Abstract

Ensuring safe drinking water is one of the top priorities in public health as waterborne diseases remain a global challenge. In China, microbial contamination in drinking water is of particular concern and comprehensive survey/monitoring of the drinking water microbiome is necessary. However, traditional culture-based microbial monitoring methods have significant limitations, and nationwide tap water survey/monitoring in China would require significant resources. Here, a cost-effective and culture-independent citizen science approach was developed to sample the microbiome in household drinking water (n = 50) from 19 provinces in China from December 2020 to August 2021, including a few opportunistic samples collected in situ right after extreme weather events such as the 2021 Henan Floods and Typhoon In-Fa Landfall. Using a protocol optimized for low-biomass samples, 22 out of 50 tap water samples were tested positive for microbial DNA. 16S rRNA gene metabarcoding were conducted on pooled samples, yielding 7,635 Amplicon Sequence Variants (ASVs), which revealed a diverse microbiome in household tap water. Alarmingly, pathogenic bacteria including *Mycobacterium* spp., *Acinetobacter* spp., and *Legionella* spp. were detected in all PCR positive samples. Despite the limited number of samples, a significant number of pathogenic species (e.g., *Salmonella enterica*) and/or toxin-producing cyanobacteria (e.g., *Microcystis* spp.) were detected in local tap water samples from Zhengzhou and Changzhou following the 2021 Henan Floods and Typhoon In-Fa Landfall, respectively. Overall, this study underscores the utility of citizen science in enhancing microbial monitoring and informs future public health strategies for water safety.

## 1 Introduction

With the growing demand for safe drinking water, microbial contamination in water resources and its related diseases is a focus of the world’s water quality control today. Numerous studies show that drinking water with microbial contamination can cause both acute and chronic damage to human health, with waterborne diseases posing a significant global health burden [1, 2]. There has been a rising incidence of waterborne human diseases such as gastrointestinal illness (e.g., diarrhea) and liver cancers which could be fatal [3–5]. The prevalence of human pathogens and toxin-producing bacteria in ambient and drinking water is a severe problem recognized globally [6]. Therefore, the scarcity of water resources that many countries including China have been facing is further exacerbated by water contamination [4, 5, 7].

A systematic review of China’s drinking water sanitation from 2007 to 2018 shows that microbial contamination in drinking water is a particular concern in China [7]. To assess the potential health risks and obtain necessary data for microbiological water safety management in China, water monitoring, which supplies a comprehensive understanding of spatiotemporal patterns in drinking water microbiome, is urgently needed, especially at the point of use [8]. China CDC (Centers for Disease Control and Prevention) at all levels sample drinking water twice a year to obtain copious water quality data [7]. Due to the vastness of China, this nationwide water monitoring requires considerable investments of capital, time, personnel, and technology [7]. Fortunately, previous research has shown that citizen science can be an effective tool to increase spatial and temporal coverage of data [9]. In the context of China, citizen science could be a cost-effective approach to supplement China’s national professional water monitoring systems [10, 11].

Citizen science can be broadly defined as a scientific approach in which the public (i.e., people who have limited knowledge and skill in the targeted field) participate in the generation of scientific knowledge [11–14], commonly in data collection [11–14]. Citizen science has a history of several centuries in western societies, particularly in the environmental domain of which the breadth is immense [10–12]. Among citizen-based environmental monitoring programs, water resources monitoring is one of the major emerging fields [10]. It is especially active in Western countries [13, 15–17] since the provision of safe drinking water is a defining aspect of a developed country [18]. The National Water Quality Monitoring Council (NWQMC) website, for example, has over 350 volunteer monitoring groups registered across the U.S. in 2018 [15]. A more recent research pointed out that citizen science will play an increasing important role in promoting freshwater research, improving public understanding of the necessity to protect aquatic ecosystems, and engaging local communities and stakeholders in freshwater resource management [19].

However, several research gaps persist. First, compared with the long history and prosperity of citizen science development in Europe and the US, little research has been done in developing countries including China [15]. This can be attributed to multiple barriers, such as the late commencement of citizen science initiatives in China, low participation levels, and issues concerning data quality control, etc. Consequently, the cooperation between Chinese scientists and the public are limited to a few citizen-based environmental projects mainly focusing on birds and plants monitoring. However, with escalating concerns over environmental issue and the growing prevalence of big data and social media in China, a new era of citizen science in China is emerging [20]. For example, a recent study conducted by Wu et al. [21] revealed that most of the citizen science projects in China aiming to improving water quality are still ongoing, indicating great potential of the citizen science approach for water monitoring in the country.

Second, a global review of citizen science projects related to water quality measurement in the past 20 years [15] shows a significant focus on chemical-physical parameters such as nutrients, water transparency, and temperature, with very few addressing waterborne pathogens. Among these waterborne pathogen assessments, only *E. coli*, an indicator organism of microbial contamination risk in water used worldwide including China [6], was targeted. However, the efficacy of current indicator organisms in representing the potential presence of pathogens in water resources is still a subject of ongoing debate [2, 6]. Furthermore, a study suggests that China should incorporate additional microorganisms as alternative contamination indicators to improve its water quality management [5]. Therefore, it is necessary to holistically assess microbial community compositions and pathogens in drinking water.

Third, traditional microbiological monitoring of drinking water generally relies on culture-based methods, such as the heterotrophic plate counts (HPC) of certain microbes [22, 23]. However, these methods can only account for a very small fraction (< 1%) of the drinking water microbiome [23–25]. To overcome this limitation, this study employed a culture-independent approach: microbial 16S rRNA gene metabarcoding. This method enabled a holistic analysis of the microbial communities in the water samples [23, 26, 27].

To the best of our knowledge, this study is one of the first to utilize a citizen science approach to sample microbiome samples from household drinking water (i.e., tap water) across different regions in China, through a simple yet standardized methodology. Following the protocol developed by this study, citizen scientists (college student volunteers) collected 50 tap water samples from households across 32 administrative regions within 18 provinces/regions of China from December 2020 to August 2021. This included several opportunistic samples collected after extreme precipitation events such as 2021 Henan Floods and Typhoon In-Fa Landfall. We successfully extracted microbial DNA from these low-biomass tap water samples, pooled all samples with positive 16S rRNA genes PCR amplification signals, and conducted high-throughput sequencing of the purified amplicons. This allowed us to characterize the total drinking water microbiome and identify potential waterborne pathogens.

## 2 Materials and Methods

### 2.1 Sample Collection

#### 2.1.1 Sites and Participants

The sampling sites (Fig 1) were determined based on the coverage area (i.e., to cover as large an area as possible) and the possibility to recruit undergraduate student volunteers from the Duke Kunshan University (DKU) community. Due to the distribution of volunteers, samples mostly came from central and eastern China (latitude: ∼22°N - 40°N; longitude: ∼100°E - 122°E), including Beijing, Shandong Province, Jiangsu Province, Guangdong Province, etc.

**Fig 1.**
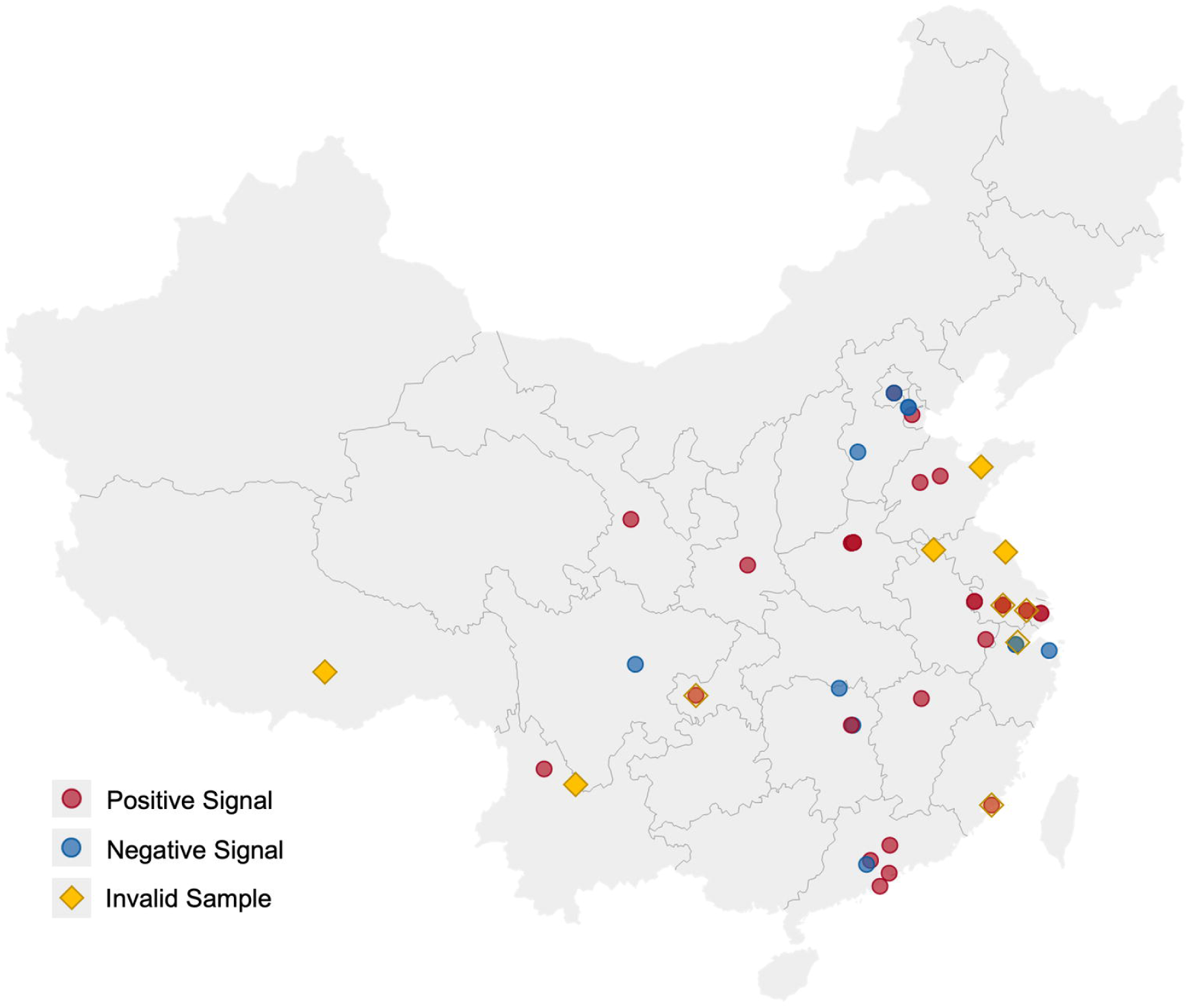
Geographical locations of sampling sites. Map showing the geographic locations of the citizen science samples and the PCR amplification results (Software: Datawrapper). Red dots denote tap water samples showing positive PCR signals, blue dots indicate negative PCR signals, and yellow diamonds represent samples that were collected but did not pass quality control. Some locations were sampled multiples times at different time points.

All student volunteers were recruited after a simple screening process. First, their relevant experiences (e.g., majors) were considered and those with natural science majors and lab experience were preferred. Besides, ideal volunteers would go home or go on a trip to any place(s) in China during the sampling period. Volunteers who met those two criteria or at least one of them were recruited through one-to-one conversation on WeChat (a popular Chinese communication app similar to WhatsApp). Communication between volunteers and investigators is a key component of this study to ensure the quality of the samples as much as possible. This is the main reason why volunteers were recruited from the DKU community – they can easily reach out to us either in person or via WeChat once any problem emerges.

#### 2.1.2 Preparation and Tool Kit

A sampling protocol was developed based on Buxton et al.’s [16] research on citizen science methods because volunteers in this study performed highly similar tasks (i.e., water sample collection and filtration) as their research. A detailed version of the protocol is provided in the “Citizen science sampling protocol and materials” (CS 1). Two innovative aspects of this protocol are (1) the easy-to-use and low-cost Corning syringe filters (instead of pumps and Sterivex filter cartridge) were adopted for water sampling, and (2) the disinfection procedure was emphasized by listing all the possible exposed objects and surface during the entire sampling process. Briefly, it is recommended to conduct the experiment on the day or at most one day before shipping the sample. Volunteers first put on the gloves and disinfect their hands as well as everything that they may touch during the operation using the disinfectant wipes (Brand: Mian Zhi Run, 75% Ethanol and RO purified water). Then, a sterilized 1 L stand-up bag with sodium thiosulfate to remove residual chlorine was unsealed and filled with 1 L tap water. A disposable sterile 50 mL syringe was used to pass the sample water across a sterile Corning syringe filter unit (CLS431229, 0.20 μm pore size, 28 mm diameter) and refilled until 1 L of water has been filtered or the filter unit has become blocked. Afterwards, a syringe of air was pushed through the filter unit to reduce the amount of residual water in the sealed unit. The filter unit was then sealed in a Ziplock bag and kept frozen in a household freezer prior to transportation to the laboratory (DKU Environmental Research Lab in Kunshan). For transportation, the protocol requires burying the filter unit sample among four to five ice packs in a Styrofoam box. Depending on the distance between the sampling site and Kunshan, along with the student’s mode of travel (i.e., same-day flights or high-speed train), samples were either delivered via express delivery service or personally carried to the laboratory by volunteers.

To further clarify the procedures and reduce variability in sample collection, an 11-minute video tutorial (720p resolution) was created, providing volunteers with a visual guide on the tools and procedures for sample collection, filtration, storage, and shipping (Fig 2 & 3 and CS 3). Furthermore, in-person demonstrations were provided at DKU Environmental Research Lab for available volunteers following the demonstration model of Willis et al. [28]. A compact sampling kit was distributed to each volunteer before their trip (Fig 2). Each kit (in a 78.7×51.2×49.2 inch Styrofoam box) contained one set of all tools mentioned in the protocol and a sampling information form (CS 1 & 2) adapted from Buxton et al.’s [16]. This form collected information such as volunteer names, sampling time, geographic coordinate of the sampling site obtained from cell phone, etc. Alongside the form, volunteers were requested to label the Ziploc bag containing the sample with their names for convenient sample tracking in the stage of data collection. Upon recipient in lab, each sample was associated with an anonymous ID number and any data with identifiers was securely eliminated, ensuring no specific identity-related information was present during data processing. The ID number of each sample consists of two parts separated by an underscore – a two-letter abbreviation of the sampling site (city) followed by a four-digit sampling date (S1 Table). For example, the sample collected in Beijing on June 20 was named as “BJ_0620” and the full names of other mentioned samples are provided in the “Non-standard abbreviations” section.

**Fig 2.**
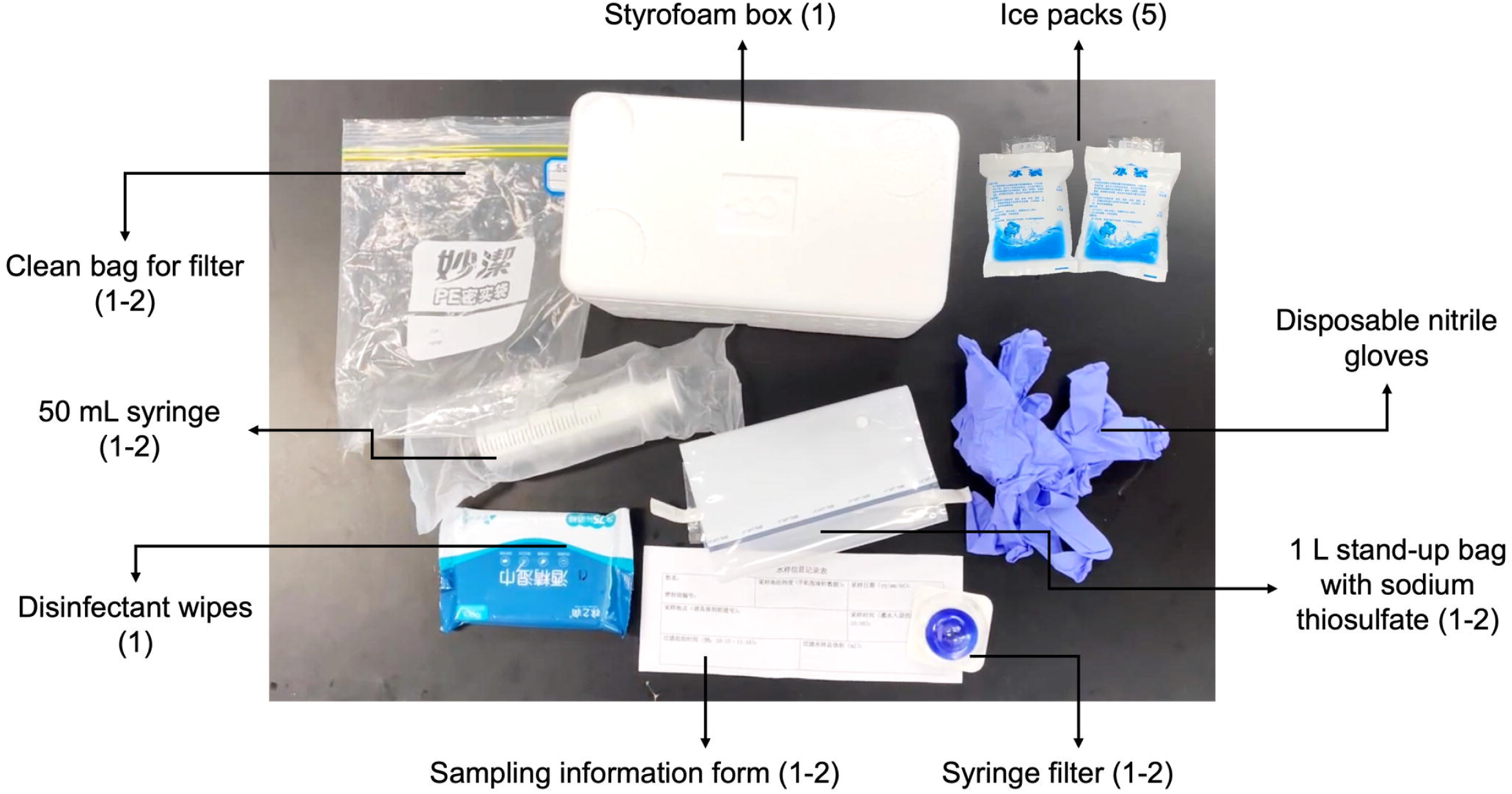
The sampling tool kit.

**Fig 3.**
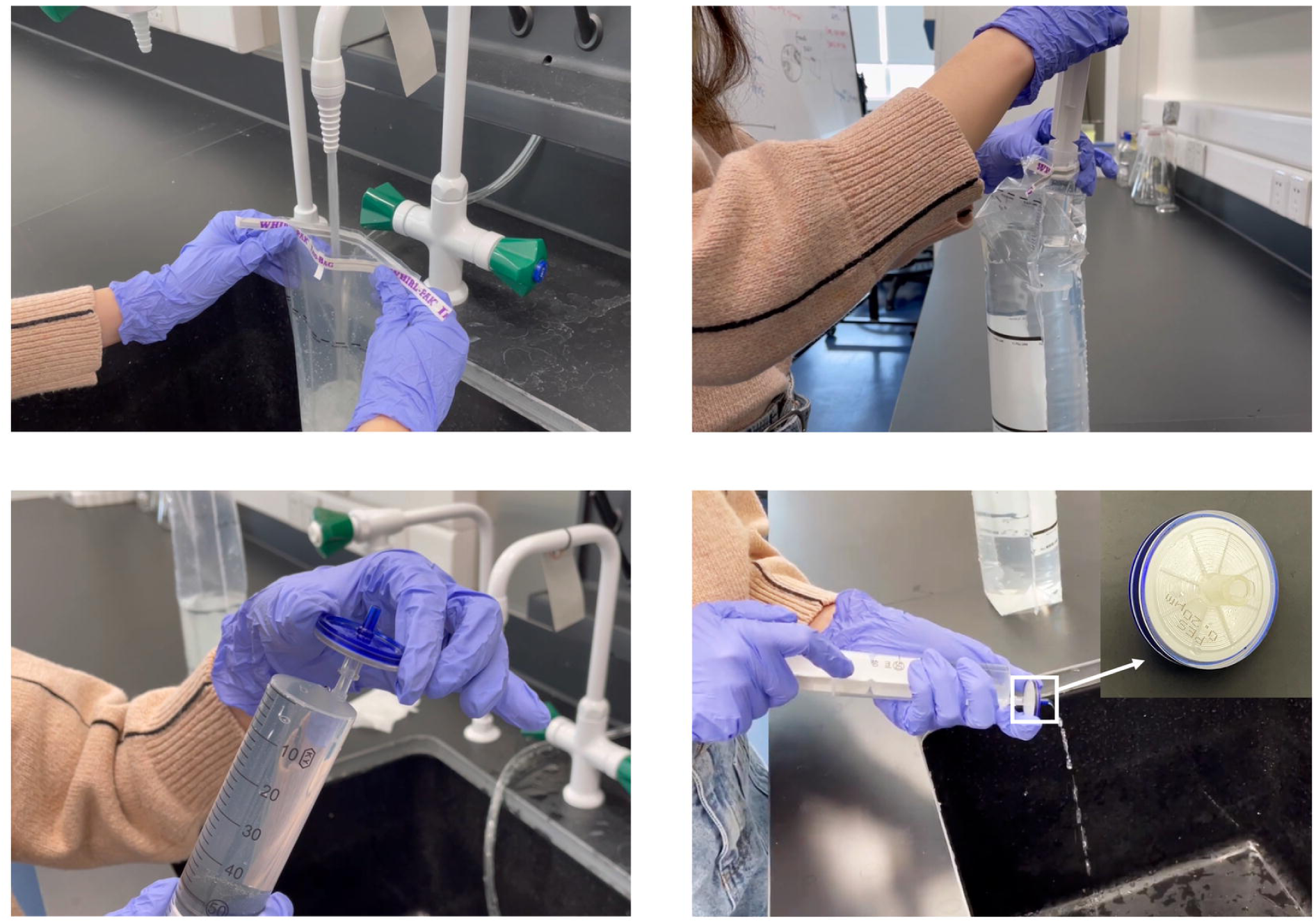
Screenshots from the demonstration video showing the sampling process and the syringe filter.

An online sharing document (Tencent Sharing Document, with English translation provided in CS 3) was sent to each volunteer via WeChat, including the brief introduction to the research project, sampling kit, and sampling protocol as well as the links to the video tutorial and the electronic sampling information form (a Qualtrics survey was created to supplement the printed form). Notably, key elements in the document were highlighted in red and a larger font size for clarity. All the materials were provided in Chinese to facilitate understanding among the recruited student volunteers who were all Chinese. The online sharing document enabled them to easily see any modifications that were made later.

Before the recruitment of volunteers, several trial runs of sampling were conducted by the authors, including sampling sites in Kunshan, Hangzhou, Changzhou, and Beijing. Along with the sampling protocol and kit, methods and conditions for sample shipping (i.e., express delivery vs. same-day bullet trains or planes) were tested and compared to identify the most effective procedures for DNA preservation.

#### 2.1.3 Sample Collection and Filtration

The formal sample collection by student volunteers was conducted from December 2020 to August 2021. At each sampling site, one to four samples were collected. For sites with more than one sample, all valid samples were included to represent the site’s microbiome. With the help of 25 volunteers, 50 household drinking water samples were collected from 32 administrative regions spanning 19 provinces/regions in China.

During this study, two extreme weather events were captured by opportunistic sampling. Tap water samples were collected by volunteers from Changzhou, Jiangsu Province and Hangzhou, Zhejiang Province before and after the landfall of Typhoon In-Fa (July 22 – 31, 2021 in China), and from Zhengzhou, Henan Province following an extremely destructive flood event (2021 Henan Floods).

Typhoon In-Fa (number 2106) was a Category 2 typhoon (SSHWS) which has been the second-wettest tropical cyclone ever recorded in China. As a tropical storm, it consecutively hit Putuo District of Zhoushan and Pinghu in Zhejiang Province on July 25 and 26 respectively. Typhoon In-Fa passed nearby Hangzhou from July 25 to 26 as a typhoon and Changzhou from July 26 to 27 as a tropical storm [29]. On the other hand, 2021 Henan Floods in Zhengzhou (July 17 – 23, 2021) was indirectly influenced by Typhoon In-Fa. From July 19 to 21, Zhengzhou suddenly encountered the most severe rainstorm of the last 50 years which has led to extremely severe urban inland inundation, floods, and landslide. In particular, the 24-hour precipitation in Zhengzhou from 8:00 on July 20 to 8:00 on July 21 was 624.1 millimeters (2.05 ft), which is only about 14.4 mm (0.57 in) lower than the average annual precipitation in Zhengzhou in the past 22 years (638.5 mm) [30]. According to the Investigation Report on 2021 Henan Floods issued by Ministry of Emergency Management of the People’s Republic of China [31], the flood impacted over 14 million individuals and killed 398 people in Henan, with the number of deaths in Zhengzhou accounting for 95.5% of the total death. Moreover, it resulted in 16 million hectares of submerged agricultural land and direct economic damages amounting to $20.69 billion, accompanied by significantly higher indirect expenses.

For the areas affected by the typhoon landfall, student volunteers were instructed to sample tap water once pre- and post-typhoon respectively within a week. As for Zhengzhou, since the sudden occurrence of the extreme rainfalls and flooding was not anticipated and predicted by the China Meteorological Administration [32], pre-flooding sampling was not arranged. However, we successfully coordinated with and supplied the sampling kits to two volunteers residing in Zhengzhou – one living in a seriously affected region and the other in a moderately affected region – during the flood. They were instructed to collect three to four tap water samples on different days throughout the week following the flood.

### 2.2 DNA Extraction, PCR Amplification, and Bacterial 16S Metabarcoding

At the DKU Environmental Research Lab, all samples were stored in a -80 freezer prior to processing. For each sample, the filter membrane was removed from the sealed syringe filter unit and cut into eight strips using a sterile razor blade on a disposable petri dish. DNA extraction was then conducted using the Qiagen DNeasy Plant Mini Kit following the manufacturer’s instructions, with the exception of an additional bead-beating step implemented to enhance extraction efficiency. In brief, the filter strips were placed into a 2 mL microcentrifuge tube where 0.2 g of 0.1 mm Zr bead and 400 microliters of lysis buffer AP-1 were added for beat beating at 2000 rpm for 5 minutes. The final extracted DNA of each sample from the kit was dissolved in 40 μL Qiagen elution buffer and stored in a -80 freezer.

For PCR amplification, the V4 region of 16S rRNA gene was amplified by the universal primer pairs 515F (5’-GTGCCAGCMGCCGCGGTAA-3’) and 805R (5’-GACTACNVGGGTATCTAAT-3’) with dual barcode index and heterogenous spacers [33, 34]. KAPA HiFi PCR Kit and the manufacturer’s protocol was adopted. All PCR reactions were performed in triplicates with 25 μL of each reaction mixture. Agarose gel electrophoresis was then performed to visualize amplicon fragments and PCR products were purified using QIAquick PCR Purification Kit (Qiagen) following the manufacturer’s instructions. The purified PCR amplicons of each sample was dissolved in 30 μL elution buffer and stored in a -80 freezer.

Finally, the concentration of the purified PCR amplicons was measured by Qubit^™^ 4 Fluorometer. Equimolar amounts of purified amplicons were pooled together and sent to *Genewiz* in Suzhou for an Illumina Miseq (250 PE) sequencing run.

### 2.3 Auxiliary Data

Previous studies found that drinking water microbiome exhibited seasonality which was correlated with temperature [35, 36]. Given the difficulties in measuring water and indoor temperature with the citizen science approach, outdoor temperature was collected as the indicator of seasonal environmental temperature change. The outdoor temperatures when collecting valid samples (n = 40) were retrieved from Weather Underground [37], an online portal that provides local weather data on an hourly basis.

In addition, to assess the potential risks of harmful bacteria detected in sampled tap water, information such as drinking water standards and microbial fact sheets was retrieved from environmental and health authorities including The United States Environmental Protection Agency (EPA) [38, 39], and World Health Organization (WHO) [40].

### 2.4 Data Analysis

#### 2.4.1 Sequence Processing

To profile the microbiome in sampled tap water, 16S rRNA gene sequences were processed by a series of bioinformatics tools. First, paired-end sequencing reads with dual indices were demultiplexed and then trimmed to remove barcodes and primers using *Cutadapt* [41]. The resulting reads were then further processed following the *DADA2(v1.16)* pipeline [42]. Specifically, using the embedded functions in *DADA2*, quality filtering was performed before merging paired-end sequencing reads. After chimera checking, Amplicon Sequence Variant (ASV) was identified, and the abundance table for each sample was constructed. Finally, the “assignTaxonomy” function in *DADA2* was used to assign taxonomy to each ASV based on Silva r138 reference database (DOI 10.5281/zenodo.4587955).

#### 2.4.2 Microbial Community Structure

The relative abundance (RA) of all ASV in each sample was calculated and the following analyses of microbial compositions were based on RA.

Apart for the samples collected after Typhoon In-Fa and 2021 Henan Floods (“post-weather samples”), the remaining samples (“normal samples”) were divided into three nearly equal groups based on the outdoor temperatures to account for potential impacts of climate and seasonality: (a) High (H): T > 25 °C; (b) Medium (M): 15 °C < T ≤ 25 °C; (c) Low (L): T ≤ 15 °C (Table 1). The mean RA of each ASV in the three temperature categories was calculated for the ASV-level analyses. The most abundant ASVs in each category are presented in the supporting material S1 Text and S1 Fig.

**Table 1.** Sampling seasons: outdoor temperature categories.

| Category | Range (°C) | Number of samples |
| --- | --- | --- |
| High | $T > 25$ | 7 |
| Median | $15 < T \leq 25$ | 7 |
| Low | $T \leq 15$ | 6 |

#### 2.4.3 Assessment of Potential Waterborne Bacterial Pathogens

The bacteria genera in the dataset containing pathogenic species were selected based on *Aquatic Pollution: An Introductory Text* [43] and “Guidelines for Drinking-water Quality (4^th^ Edition)” by WHO [40]. Subsequently, the taxonomy of the resulting ASVs was further validated using the BLAST+ tool [44] against the NCBI database during February 2022. Based on our criteria, only BLAST results with percent identity (p-ident) > 97% [45], expect value (E-value) < 10e-100 [46], and query cover equals 100% were considered reliable results.

Subsequently, all potential pathogenic genera/species were grouped into two categories based on their occurrence in the drinking water samples. Those detected in more than 30% of all samples (i.e., 7 samples) were categorized as “common pathogens” while the rest were referred to as “rare pathogens.”

#### 2.4.4 Statistical Analyses

All statistical analyses were performed in R (4.1.2 and 4.2.3) and a p-value threshold of 0.05 was considered significant.

Fisher’s exact test [47] was conducted to determine whether there is a significant relationship between PCR signals (i.e., positive or negative) and average annual rainfall (AAR) (humid regions: > 800 mm, non-humid regions: < 800 mm).

Alpha-and beta-diversities were calculated at the ASV level to further describe the microbial communities in the samples. Alpha diversity refers to diversity (richness or evenness) within an ecosystem at the local small scale, while beta diversity measures the amount of differentiation between different ecosystems or local species communities [48]. For alpha-diversity of tap water microbial communities, each library was resampled in equal depth, and Chao1, Fisher, Shannon, and Simpson diversity indices were then calculated from observed read counts of ASVs using the “Phyloseq” package (version 1.38.0) in R [49]. The Shannon and Simpson indices including both ASV richness and evenness were computed because of their reduced sensitivity to differences in sample depth [50, 51]. To calculate beta-diversity, a principal coordinates analysis (PCoA) was performed based on the distance matrix of Bray-Curtis dissimilarity to visualize the ordination among the samples by *plot_ordination()*. To test the significant differences among groups of tap water samples, permutational multivariate analysis of variance (PERMANOVA) and a post-hoc pairwise Adonis test were performed with adonis2 function [52] and *pairwise.adonis()* function [53] respectively. To check if within-group variation is confused with among-group variation [54], a permutation test for homogeneity of multivariate dispersions (PERMDISP) [55] was performed with the *betadisper* function. In PERMANOVA and PERMDISP, the number of permutations was set to 999.

To analyze the compositions and influencing factors of waterborne pathogens in the tap water, several correlation tests based on Spearman’s rank correlation coefficients were conducted. First, Mantel test [56] was conducted to examine correlations between total microbial communities (top 100 ASVs), potential pathogens, and outdoor temperatures based on Bray-Curtis dissimilarity, Euclidean Distance, and Haversine Distance matrices (permutations = 9999). Second, linear regression fits between outdoor temperatures and RAs of total potential pathogens were examined and visualized with *ggscatter()* in the “ggpubr” package. Third, linear correlations between RAs of potential pathogens, RAs of total microbial communities (Top10 ASVs), and alpha diversity of total microbial communities were assessed with *cor()*. The corresponding p values and confidence intervals were computed with *cor.mtest()*. The matrix with significance level codes was visualized with *corrplot.mixed()*.

## 3 Results and Discussion

### 3.1 Sample Validity and PCR Signals

Out of the 50 drinking water samples, 40 passed our quality control and they were collected from 28 administrative regions across China (Table S1). 10 samples were excluded from the study because of either (1) lab processing errors, or (2) improper handling during shipping and/or storage, which may have compromised DNA quality. Among the 40 valid samples, 29 samples showed positive PCR signals which were collected from 16 cities in 10 provinces, 4 municipalities, and Macau (Fig 1). Fisher’s exact test (S2 Table) reveals that no significant difference in PCR signals existed between the two rainfall groups (P = 0.69). Interestingly, all the three tap water samples collected in Zhejiang Province, including the post-typhoon Hangzhou sample showed no PCR signal. This might be due to the high quality of source water in Zhejiang Province [57] and effective drinking water treatment in those two developed cities (Hangzhou and Ningbo). Alternatively, factors such as new/clean plumbing systems, suitable plumbing materials that do not support the growth of microorganisms, and shorter water stagnation time in the plumbing could also account for the negative PCR results [35, 58, 59].

### 3.2 Citizen Science Sampling Approach

This study is among the few studies on tap water microbiome via citizen science which can serve as a proof of concept for national-scale microbiological monitoring of tap water using citizen science and demonstrate its competitive advantages compared to non-citizen science sampling method.

Firstly, the study illustrates the cost-effectiveness and extensive coverage across China of the approach. With decentralized volunteer participation and affordable sampling kits, the sampling sites spanned 32 regions and various seasons, ensuring spatial and temporal diversity. It is worth mentioning that due to the adoption of Corning® syringe filter (∼$2.5–4/unit) [60], the cost per sampling kit was under $7, which was around 1 – 1.5 times cheaper than using the traditional MilliporeSigma® Sterivex filter (∼$8–13/unit) for aquatic microbiome sampling [61]. Our DNA extraction protocol was optimized for this low-cost filter. In addition, this citizen science sampling approach significantly reduces travel and personnel expenses. Although the volunteers’ sample collection lacked strict oversight, the high validity of 40 out of 50 drinking water samples passing quality control and the above consistency in results supports the method’s applicability and reliability. Furthermore, due to the flexibility of this sampling approach, we conducted timely opportunistic sampling of extreme weather events – specifically, Typhoon In-Fa and 2021 Henan Floods – by leveraging local volunteers in Hangzhou, Changzhou, and Zhengzhou. This citizen science approach proved to be particularly fast responding and effective given the limited funding and the urgent nature of these events. Additionally, the sample storage tests (S3 Text) confirmed that maintaining samples for 7 days at -18°C did not generate false positives in the absence of bacteria during collection, ensuring accurate pathogen identification and reliability of using household freezer for storage, thereby enhancing the flexibility and feasibility of the citizen science sampling approach. Moreover, the citizen science approach aimed to bridge the gap between the scientific community and the public, raising awareness of pathogen pollution in drinking water and educating the public on health protection measures.

### 3.3 Sequence Reads, ASVs, and Taxonomy Classification

The DNA of microorganisms in 29 household drinking water samples were successfully extracted, PCR amplified with 16S primers and submitted for Illumina sequencing. However, following the demultiplexing process, five samples did not yield significant reads (all < 50 per library) and were excluded from the dataset, leading to a loss of two sampling sites (Shenzhen and Xiamen). In addition, two samples collected after the 2021 Henan Floods were excluded after the quality filtering and chimera checking because of relatively low reads (<3000 per library) compared to others. As a result, the 16S rRNA gene amplicon dataset in this study contained 22 samples after the preliminary processing (S1 Table).

In DADA2 quality filtering, proportions of output and input reads of 13 (59.1%) samples were lower than 50%, indicating low quality of raw DNA reads. This may be due to 1) potential degradation of DNA samples during sampling, shipping, or DNA processing; 2) residual DNA from dead bacteria cells, which are harmless to humans. After quality filtering and chimera removing, sequencing reads per sample ranged from 9,794 to 150,656, averaging at 60,884.

From the resulted dataset, 7635 ASVs were identified. According to the DADA2 taxonomy classification based on Silva r138 reference database, 100%, 99.5%, 96.8%, 92.3%, 81.8%, and 59.4% of the sequencing reads could be assigned to kingdom, phylum, class, order, family, and genus level respectively. At the kingdom level, 99.9% of the reads were assigned to domains of bacteria (1,294,678 reads) and archaea (43,816 reads), while the other 0.1% of the sequences belonged to eukaryotes (739 reads).

### 3.4 Microbial Community Structure

#### 3.4.1 Compositions of Total Microbial Communities

52 microbial phyla including 46 bacterial phyla and 6 archaeal phyla were detected in the household drinking water samples (n = 22). In all samples, 9 bacterial phyla and 1 archaeal phylum accounted for over 90.7% of total taxonomically assigned reads at phylum level (Table 2). All the ten phyla are both abundant and prevalent (i.e., occur in 91-100% of all samples), indicating a relatively even microbial distribution pattern in the samples at the Phylum level. The five most dominant bacterial phyla were *Proteobacteria* (mean percentage ± SD: 55.0% + 19.8%), *Planctomycetota* (10.5% + 7.9%), *Acidobacteriota* (7.0% + 6.0%), *Actinobacteria* (5.9% + 6.6%), and *Cyanobacteria* (4.7% + 3.5%) , comparable to a previous study in China [57]. Three of them were reported tolerant to drinking water treatment and distribution processes except for *Planctomycetota* and *Acidobacteriota* [57]. Interestingly, this bacterial composition differs from the primary bacterial assemblages revealed by several highly-cited studies conducted in other countries including the U.S. and Portugal, which predominantly feature *Proteobacteria*, *Actinobacteria*, and *Bacteroidetes* [62–64]. This implies a unique microbiome intrinsic to China’s drinking water systems.

**Table 2.** Number of sequences, ASVs, and genera for the top 10 phyla in the water samples.

| Phylum | Sequence <sup>a</sup> | ASV | Genus |
| --- | --- | --- | --- |
| *Proteobacteria* | 697642 (52.3%) | 2693 | 281 |
| *Planctomycetota* | 164487 (12.3%) | 998 | 23 |
| *Actinobacteriota* | 82262 (6.2%) | 279 | 56 |
| *Acidobacteriota* | 77415 (5.8%) | 296 | 17 |
| *Cyanobacteria* | 72219 (5.4%) | 402 | 25 |
| *Crenarchaeota* | 43624 (3.3%) | 31 | 5 |
| *Bacteroidota* | 38869 (2.9%) | 603 | 61 |
| *Verrucomicrobiota* | 33769 (2.5%) | 411 | 27 |
| *Bdellovibrionota* | 31056 (2.3%) | 400 | 8 |
| *Desulfobacterota* | 21608 (1.6%) | 59 | 11 |
| Others | 70371 (5.3%) | 1209 | 93 |
| N.A. | 6133 (0.5%) | 254 | / |

Those three represent the coverage, diversity, and the genus spectrum of microbial community in the water samples respectively. <sup>a</sup> The percentage of sequences was the phylum sequences in the total of assigned sequences at the phylum level which was 1,333,322.

The phylum-level taxonomic composition for each sample is detailed in Fig 4 and S3 Table. In this study, *Proteobacteria* (class α*-* and γ*-Proteobacteria*) was the most predominant phylum in 20 samples and the second most in others. It accounts for 29.9 – 99.0% of the reads in each sample, with the highest relative abundance detected in Huizhou (HZ_0219). Among all the samples, *Sphingomonas* was the most abundant genus of *Proteobacteria*. This result was consistent with the previous study [57] that found *Proteobacteria* to be dominant in tap water collected mostly from central and eastern China, and the genus *Sphingomonas* grew during chlorination [65] or monochloramine treatment [66].

**Fig 4.**
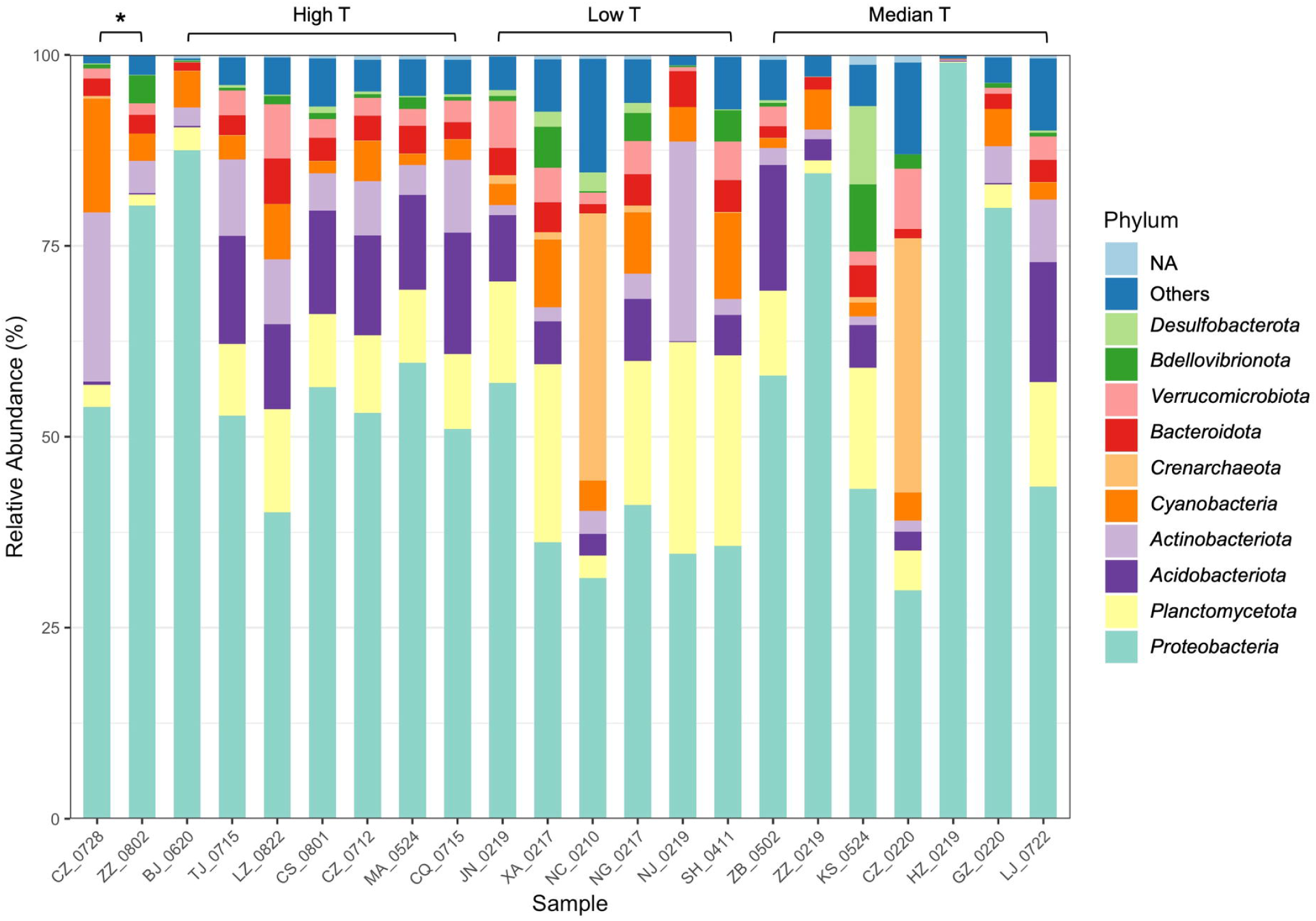
Taxonomic composition and relative abundance of microbiota in the sampled household drinking water in China at the phylum level. “*” denotes the samples collected after extreme rainfall events. Normal samples are grouped based on the outdoor temperature. Only the top 10 phyla (1-10: bottom to top in the legend) and not assigned (NA) ones are shown.

However, two samples CZ_0220 (Changzhou, Feb. 20) and NC_0210 (Nanchang, Feb.10) were dominated by *Crenarchaeota*, a common archaeal phylum. Specifically, *Crenarchaeota* accounted for 33.3% and 35.0% with the genus *Candidatus Nitrosotenuis* (32.0%) and *Candidatus Nitrosotalea* (34.9%) being most abundant in samples CZ_0220 and NC_0210 respectively. This showed that in addition to a variety of bacteria, archaea can also grow in tap water, which could be supported by the studies which detected the archaeal phylum *Crenarchaeota* in drinking water distribution systems or drinking water-related environments [24, 67–70]. In particular, Inkinen et al. [67] found a high abundance of archaeal reads from the genus *Candidatus Nitrosotenuis* and *Candidatus Nitrosotalea* in drinking water distribution systems supplying non-disinfected waters. This suggests that the disinfection processes of samples CZ_0220 and NC_0210 may be less effective compared to others.

Interestingly, compared to the winter Changzhou sample CZ_0220, the pre-typhoon CZ_0712 and post-typhoon samples CZ_0728 collected from the same city household in July were much more similar in overall composition at the phylum level, indicating a seasonal effect. However, the post-typhoon sample CZ_0728 exhibited higher *Actinobacteria* (increased from 2.2% to 7.1%) and *Cyanobacteria* (increased from 5.2% to 14.9%), which are the phyla containing potential waterborne pathogens and the species that produce cyanotoxins, respectively. Elevated levels of the pathogen *Mycobacterium* spp. (more details in Section 3.5) as well as toxin-producing *Cyanobacteria* spp. were observed in post-typhoon sample CZ_0728. Specifically for cyanobacteria, *Microcystis* spp. were higher in RA while *Cylindrospermopsis* sp. and *Dolichospermum* sp. appeared after the typhoon event. Other toxic species of *Cyanobacteria*, including *Aphanizomenon* sp. and *Anabaena* sp., were detected in normal samples collected from Shanghai, Lanzhou, Xi’an, etc. Many *Cyanobacteria* spp. from those genera can produce a variety of cyanotoxins such as Microcystins and Cylindrospermopsin, which can cause liver and kidney damage and have potential carcinogenicity [39]. Similarly, a substantial rise in the RA of *Cyanobacteria* was reported in treated water samples collected from a drinking water treatment plant in Jiangsu Province after Typhoon Lekima in August 2019 (P < 0.05) [71]. In this study, although remained detectable, the RA of *Cyanobacteria* in post-typhoon sample CZ_0728 decreased to the pre-typhoon level by the third day after the typhoon event.

#### 3.4.2 Beta-Diversities

The beta diversity of the total microbial communities in sampled tap water is shown in Fig 5 (n = 22), S2 Text (n = 20), and S2 Fig (n = 20). According to Fig 5, Principal Axis 1, 2, and 3 for PCoA (Bray-Curtis dissimilarity) represent 20.4%, 14.5%, and 8.1% of the variation among the samples respectively. The PCoA ordination illustrates that despite the different sampling sites, the samples in High T category clustered closely except BJ_0620 which was greatly dominated by *Proteobacteria* (87.5%). In contrast, samples in the other two categories, especially Medium T exhibited dispersive distributions. According to PERMANOVA and PERMDISP (S4 Table), the influence of outdoor temperatures was statistically significant (R^2^ = 0.24, adjusted *P* = 0.012 between High T and Low T), and the differentiation was not due to differences in group dispersions (*P* = 0.17). In addition, a Mantel test (S5 Table) indicates a strong correlation between microbial community structures and outdoor temperatures (Spearman’s rho = 0.898, P = 0.0001).

**Fig 5.**
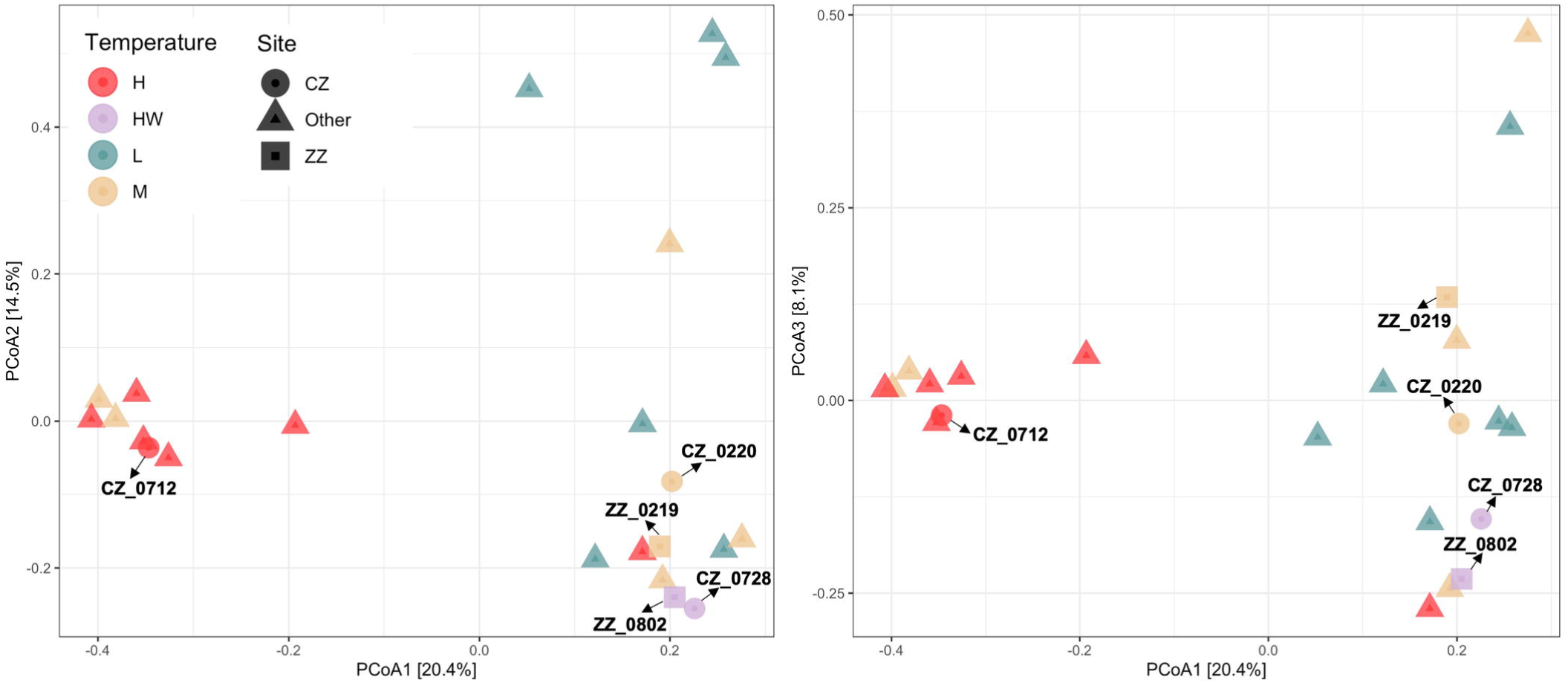
Principal coordinates analysis (PCoA). Based on the distance matrix of Bray-Curtis dissimilarity of microbial profiles (ASV-based) among all samples (n = 22). HW: the weather samples collected in summer; CZ: samples collected in the same household in Changzhou; ZZ: samples collected in two nearby households in Zhengzhou.

Research spanning China, the Netherlands, and the U.S. has consistently found that temporal temperature variation, primarily driven by seasonal changes, is a controlling factor for the tap water microbiome [35, 72, 73]. Complementing this, another study in China revealed that increasing air temperature through indoor heating could lead to dramatic changes in the composition of the bacterial community in overnight stagnant tap water [74]. These findings challenge the previous discovery by Han et al. that only a weak correlation was found between air temperature and tap water bacterial community in China (Spearman’s rho = 0.088, P = 0.092) [57]. This might be caused by differences in sampling year, sites, and geography coverage. Notable, the source water microbiome could also exert a substantive influence on tap water microbiome [57]. Beyond the citizen science approach, more comprehensive metadata collection associated with microbiome sampling, as well as well-controlled studies systematically covering spatial and temporal variations, are needed to reveal the factors shaping the drinking water microbiome.

### 3.5 Potential Pathogenic Bacteria in Drinking Water Microbiome

In total, six bacteria genera containing pathogenic species and three pathogenic species were detected in all the PCR positive samples (n = 22) (Table 3). Five genera and one species that occurred in more than 30% of the samples (i.e., 7 samples) were categorized as common pathogens while the rest were grouped into rare pathogens (S6 Table). It is worth mentioning that the overall pattern of pathogens in the normal samples still holds when the two post-weather samples were included. This finding suggests that pathogen contamination in tap water could be a widespread phenomenon [35, 75].

**Table 3.** Occurrence and mean relative abundance (RA) of major potential pathogens.

| Pathogen Species | Occurrence | Mean RA |
| --- | --- | --- |
| *Mycobacterium* spp. | 22 (100.0%) | 2.67E-02 |
| *Acinetobacter* spp. | 22 (100.0%) | 1.43E-02 |
| *Legionella* spp. | 22 (100%) | 4.59E-03 |
| *Brevundimonas* spp. | 20 (90.9%) | 6.12E-03 |
| *Leptospira* spp. | 11 (50.0%) | 2.04E-04 |
| *Escherichia coli* | 8 (36.4%) | 5.80E-04 |
| *Bacillus* spp. | 3 (13.6%) | 2.01E-05 |
| *Aeromonas hydrophila* | 2 (9.1%) | 4.76E-05 |
| *Salmonella enterica* | 1 (4.5%) | 1.79E-04 |

The distribution of these potential pathogens within each water sample are detailed in Fig 6 and the BLAST+ results of the potential pathogenic ASVs are provided in S7 Table. Common pathogens’ mean relative abundance (RA) in all tap water samples ranged from 0.02% to 2.67% (normal samples: 0.02% - 1.95%) while that of rare pathogens was extremely low (S10 Table). *Mycobacterium* spp. (mean RA 2.67%, including rainfall events)*, Acinetobacter* spp. (1.43%), and *Legionella spp.* (0.46%) occurred in all the samples while *Leptospira* spp. (0.02%) were found in half of the samples. Notably, *Escherichia coli.* ASVs (0.06%) (E-value = 6e-132, Percent identity = 100%) were detected in 36.4% of the samples. This is alarming since *E. coli* O157:H7 is the most common pathogenic strain that causes severe gastrointestinal (GI) illness in humans [76]. In addition, two *Brevundimonas* species, *B. vesicularis* and *B. diminuta*, are particularly considered emerging global opportunistic pathogens [77] were found in most of the samples (90.9%). Compared to normal samples, three more pathogenic bacteria species were detected in the post-weather samples, including *Salmonella enterica*, *Brevundimonas diminuta,* and *Aeromonas hydrophila*.

**Fig 6.**
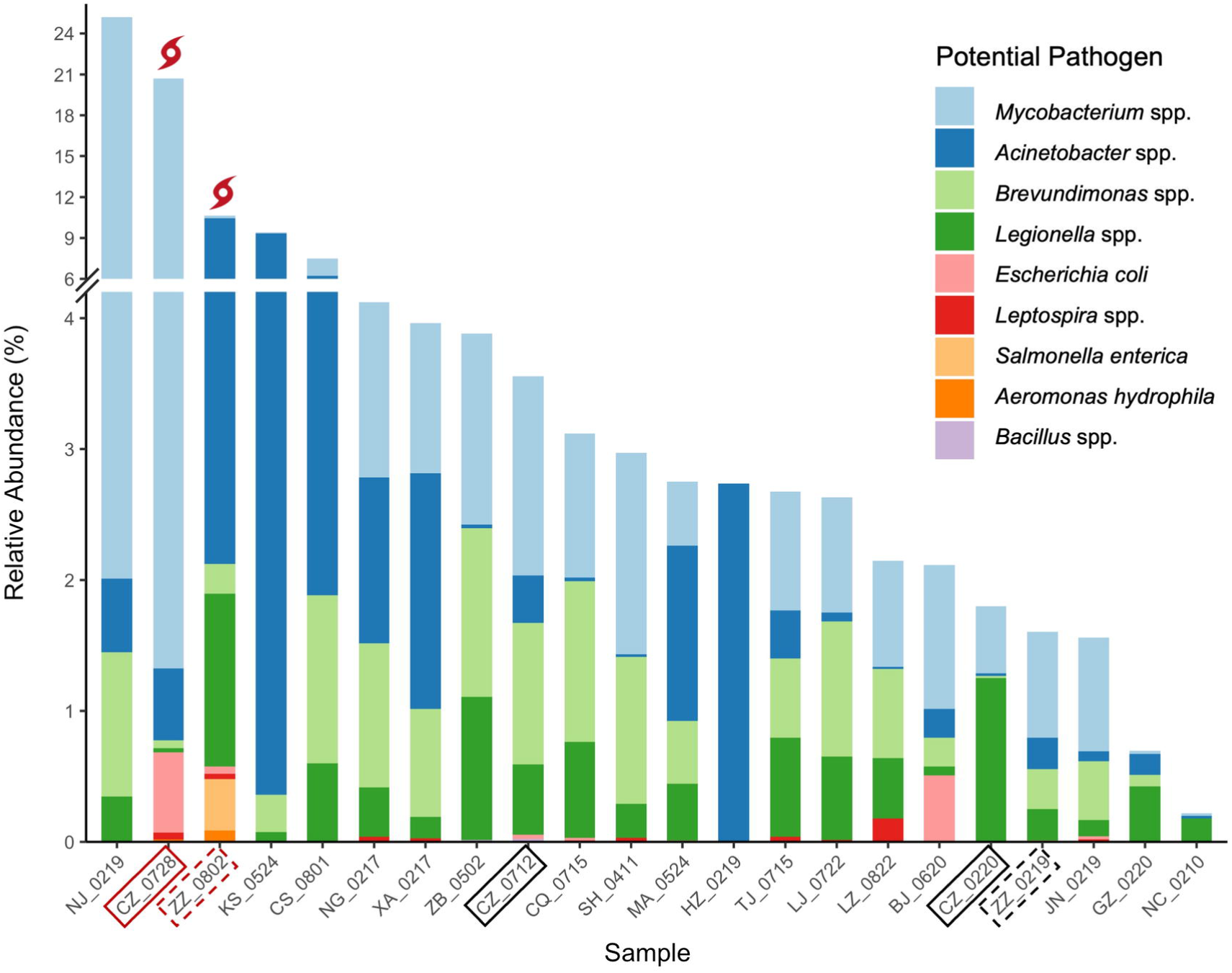
Relative abundance of potential bacterial pathogens. Pathogens in the figure legend are in the descending order of mean RA (from top to bottom). Samples are in the descending order of total RA of all pathogens in each sample (from left to right). Pre- and post-weather samples are labeled in black boxes and red boxes, respectively. Changzhou samples (typhoon) are in solid boxes, and Zhengzhou samples (floods) are in dashed boxes.

A summary of representative potential pathogens in drinking water systems and their related diseases is provided in S8 Table [1]. The discussions of several representative pathogen species for science communication purposes as well as the potential environmental drivers of waterborne pathogens are provided in S4 Text, S5 Text, and S3 Fig.

*E. coli,* a common fecal indicator bacteria [40], was widely detected by this study. The most polluted tap water samples are CZ_0728 (RA: 0.61%), BJ_0620 (0.51%), ZZ_0802 (0.05%), CZ_0712 (0.03%), and JN_0219 (0.02%). The elevated RA of *E. coli* in the two post-weather samples suggests that the contamination might be related to the extreme rainfall events. Notably, the proportion of *E. coli* in BJ_0620 was dramatically higher than that of other normal samples. It is worth mentioning that the traditional *E. coli* tests can be a generally reliable indicator of enteropathogenic serotypes in drinking water, but potential viable while non-culturable *E. coli* cells could result in underestimations of the actual water contamination [78]. Therefore, it is recommended to use PCR or quantitative PCR (qPCR) methods for the monitoring of *E. coli* [79].

Additionally, the RA of total *Legionella* spp. was most abundant in ZZ_0802 (1.32%), CZ_0220 (1.25%), and ZB_0502 (1.09%). Almost all species in the genera *Legionella* are thought to be potential human pathogens, but *L. pneumophila* (on Contaminant Candidate List 5 - CCL 5) is the main cause of Legionnaires’ disease (pneumonia) and Pontiac fever (a milder infection) [38, 40]. Potential *L. pneumophila* ASVs was detected in 22.7% (5) of the tap water samples (3 from Low T, 1 from Median and High T respectively). Among those samples, the highest *L. pneumophila* RA was observed in ZZ_0219 (RA: 0.237%), followed by MA_0524 (0.035%), NC_0210 (0.019%), XA_0217 (0.011%), and SH_0411 (0.010%). Moreover, some other pathogenic species such as *L. oakridgensis* and *L. maceachernii* occurred in TJ_0715, LZ_0822, XA_0217, LJ_0722, and ZZ_0802.

*Salmonella* spp., a highly pathogenic bacteria, was detected in one sample despite its known susceptibility to disinfection. Notably, *Salmonella enterica* (ASV 3082, RA: 0.39%) was observed in the post-flood tap water microbiome from Zhengzhou (ZZ_0802). However, the potential health risks remains uncertain, as the severity of the disease depends on the serotype and host factors of *Salmonella* [40]. Nonetheless, the presence of *Salmonella enterica* after 2021 Henan Floods indicates potential fecal contamination in the household drinking water after an extreme weather event.

Spearman’s correlations between potential pathogens and total microbiome alpha diversity indexes are shown in Fig 7. When the impacts of extreme rainfall events were not considered (n = 20), the RA of *Brevundimonas* spp. positively correlated with multiple pathogens and alpha diversity indexes, especially with *Mycobacterium* spp. (r = 0.84, P < 0.001) and the total RA of potential pathogens (r = 0.67, P < 0.001). This suggests *Brevundimonas* spp. to be a potential good candidate for indicator organisms. Notably, a significant correlation between the low alpha diversity of the microbial community and the samples positive for the presence of *E. coli* (Chao1: r = -0.24, P < 0.05; Shannon: r = -0.21, P < 0.05) was observed. Similarly, Chopyk et al. [80] found the same pattern in their pre-harvest cattle hide samples. Therefore, it is worth understanding the relationship between indigenous water microbiomes and pathogenic *E. coli*, which might contribute to the control/prevention strategies for EHEC in tap water as well as indicators for risk assessment.

**Fig 7.**
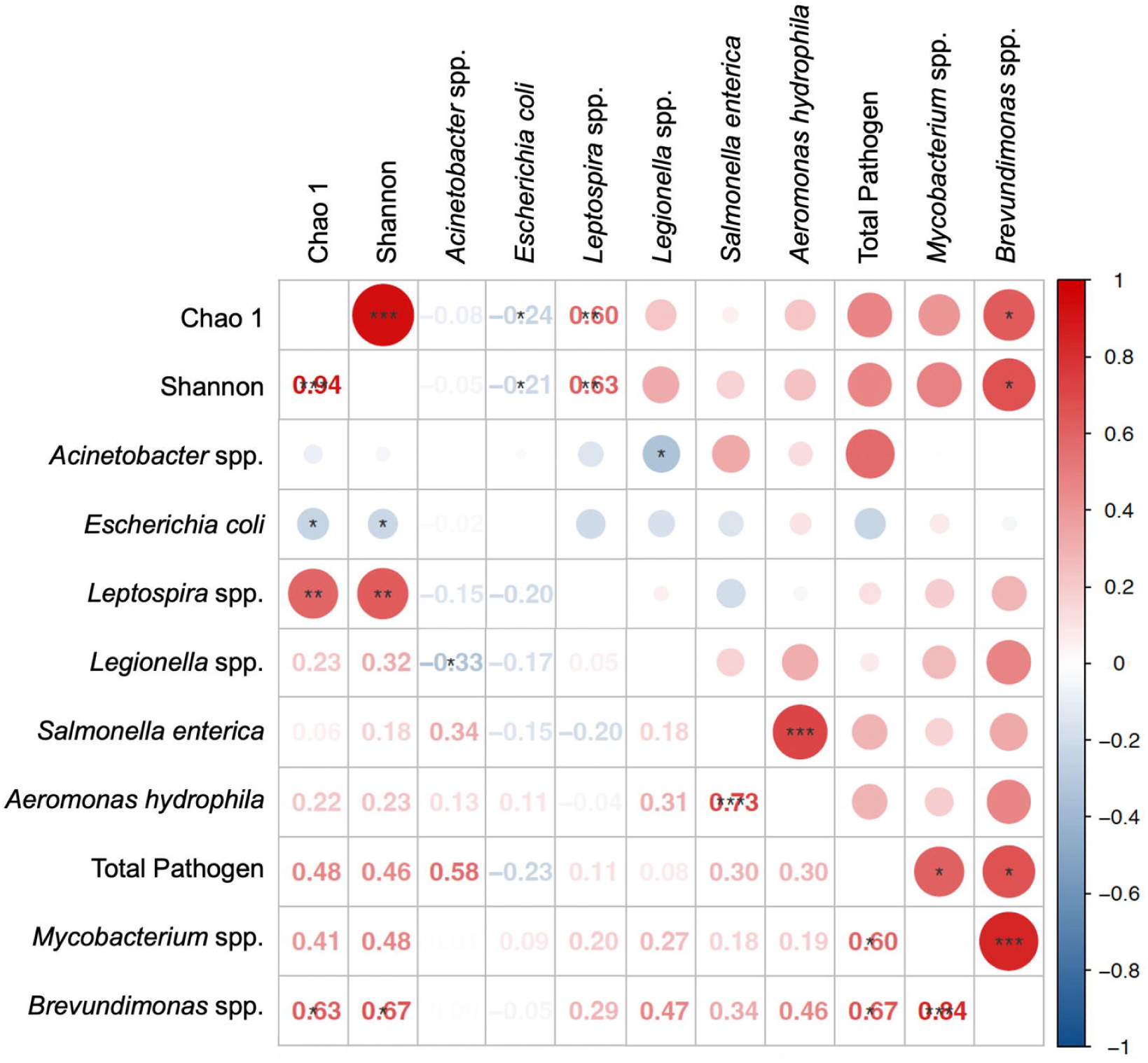
Spearman’s correlation of potential pathogens detected in normal samples (n = 20). 1-3 asterisk(s) denote the significance level of 0.05, 0.01, and 0.001, respectively.

### 3.6 Limitations and Future Work

The current sampling procedure exhibits several limitations and could be improved in the future. First, to further minimize bias during the sample collection process, closer supervision of the volunteers (e.g., through cell phone video recording) and duplicate sample collection from the same location are needed. Second, systematic time-series sampling in the same location is necessary to understand temporal microbial patterns. Additionally, to pinpoint contamination sources, future studies could sample from water treatment system effluents to discern if contamination stems from treatment or pipeline issues. Moreover, collecting a comprehensive set of metadata (e.g. water temperature, nutrients, pH, etc.) associated with the microbiome sampling will help reveal the environmental factors shaping the drinking water microbiome.

To improve the success rate of sample collection, it is important to implement stricter controls on shipping temperatures and to collaborate with shipping companies. This collaboration will help reduce sample transportation costs, a critical factor in expanding the citizen science outreach beyond university affiliates. Meanwhile, developing an online platform could be helpful to disseminate the research results to the public, thereby promoting science education and citizen engagement.

## 4 Conclusions

Ensuring household drinking water safety is vital for public health due to the risks associated with microbial contamination. Combining citizen science sampling and culture-independent metabarcoding, this proof-of-concept study provided profiles of tap water microbiome and waterborne pathogens from various locations in China. This method, which extends beyond basic water collection and observation (e.g., water transparency), suggests that well-structured collaborations between professional agencies and citizen science can effectively monitor water quality on a broad scale.

In this study, a total of 7,635 prokaryotic ASVs were detected from 40 household drinking water samples from 28 cities of 18 provinces/regions in China. Despite the limited number of samples, extreme weather events, such as typhoons and floods, may exacerbate the presence of pathogens (e.g., *Escherichia coli* and *Salmonella* spp.) as well as toxin-producing cyanobacteria *Microcystis* in local tap water. This is particularly concerning in the current context of climate change, which may increase the frequency and intensity of such conditions. Additionally, this study highlights the valuable role that citizen science can play in advancing our understanding of environmental health risks and shaping public health policy.

## Data Availability

The datasets and sampling protocol (CS) for this study will be deposited to NCBI and Zenodo (10.5281/zenodo.8012153) upon publication. The codes for data analysis are available upon reasonable request to the authors.

https://doi.org/10.5281/zenodo.8012153

## Acknowledgements

First, we would like to thank Xiuwen Li for her support in sampling and data management. In addition, we are grateful to Shuai Gu and Yuchen Meng for their help with sample processing. Last but not the least, we wish to extend our special thanks to all the student volunteers who helped collect tap water samples across China and made this citizen science project possible.

## Non-standard abbreviations

RA: Relative abundance
CS: Citizen science sampling protocol and materials
NC: Nanchang, Jiangxi
XA: Xi’an, Shanxi
NJ: Nanjing, Jiangsu
HZ: Huizhou, Guangdong
JN: Jinan, Shandong
ZZ: Zhengzhou, Henan
CZ: Changzhou, Jiangsu
SH: Shanghai
ZB: Zibo, Shandong
MA: Macau
BJ: Beijing
TJ: Tianjin
CQ: Chongqing
LJ: Lijiang, Yunnan
LZ: Lanzhou, Gansu

