## Supplementary material for "Mapping total microbial communities and waterborne pathogens in household drinking water in China by citizen science and metabarcoding": CS_Citizen science sampling protocol and materials.docx

### 1 Sampling protocol

1. Turn on the compass and positioning service of your mobile phone, record the latitude and longitude of the sampling site and take a screenshot. On the sampling information form, fill in the name, location, and date of sampling.

2. Several time points should be recorded during the whole filtration process: 1) sampling time (that is, the time when water is poured into the bag); 2) the start of filtration; 3) the end time of filtration.

3. There are four ice packs in the sampling kit. Fill the ice packs with tap water and store them in the freezer.

4. Before filtering water samples, put on gloves and use disinfectant wipes to disinfect gloves and the surface of other equipment, such as tabletop, sampling bottle, syringe, and the filter.

5. Take out the 1L stand-up bag and unseal it along the dotted line. In order to facilitate pumping, fill the bag as much water as possible, but do not exceed the fold line, otherwise the bag will not stand stably. After the stand-up bag stands stably, record the time on the information form.

6. Take out the 50 mL syringe. Before ripping the filter wrapper, first stick some transparent tape on the opening of the package. When ripping the package, rip slowly until the filter can be taken out and avoid ripping the package completely. After taking out the filter, immediately seal the package with transparent tape. Do not seal the package too tightly since you will open it again to put the filter back after filtering. The purpose of sealing the package is to keep dust and bacteria out.

7. Next, use the syringe to extract slightly more than 50mL of water from the stand-up bag. If there are small bubbles in the syringe, place the syringe vertically with the head pointing upwards (make sure the bubbles are facing the pumping hole), push the piston to drain air, and drain excess liquid against the pool until the water column is at the 50 mL mark line.

8. Record the starting time of filtration on the information form and connect the transparent end of the filter to the syringe. Note that when handling the filter, avoid touching the interface at both ends and try to hold the filter by only holding the edge of the circular plastic ring. Next, push the piston to start filtering (it is normal to feel the damping), be careful to push the piston to the end, and then grab the edge of the circular plastic ring to remove the filter.

9. Repeat the pumping and filtration process until a total of 1L of water passes through the filter (ideally it will be 20 times), or until the syringe cannot be pushed. Then record the total volume of filtered water on the information form.

10. Remove the filter, use the syringe to pump a tube of air, connect to the filter again, and push the air into the filter to reduce the amount of residual water in it. You will feel strong damping, so just try to push as much air as you can into the filter until you cannot push it anymore.

11. Record the filtration termination time on the information form. Remove the filter, place the filter into the filter package, and seal it with the tape tightly.

12. Syringes, disinfection wipes and gloves do not need to be recycled and can be dumped after sampling. Take a photo of the information form. Put the filter and the information form into a new Ziploc bag we provide in the kit and seal the Ziploc bag. Write your name on the second line of the label on the bag and the date of filtration on the third line. Store the Ziploc bag in the freezer (below -16°C).

13. Fill out the electronic Qualtrics survey based on the photo of the information form.

14. Take out the filter and ice packs from the freezer when you are ready to transport them back to our lab. Open the Ziploc bag and press the bag with your hand to expel air from the bag. After the air is mostly expelled, seal the Ziploc bag tightly. Then fold the sealed bag as shown in the video and seal it with tape.

15. Put all the ice packs (with insulation materials such as bubble pouch is better) and the Ziplocs bag into a white foam box and seal the foam box with adhesive tape. If it is not convenient to take the foam box with you when you return to campus (this method is recommended for being faster), you can make an appointment with SF Express Fresh Delivery Express service to send it back to campus with the fastest delivery option and contact us for reimbursement.

### 2 Printed sampling information form

| Name: | GPS (Longitude, Latitude): | | Sampling Date (yy/mm/dd): |
| --- | --- | --- | --- |
| Sample ID (leave blank): |  |  |  |
| Sampling Site: | | | Sampling Time (e.g., 10:00): |
| Filtering Time (e.g., 10:10–10:40): | | Total Volume of Filtered Sample (ml): | |

### 3 White book of tap water sampling and filtration via citizen science approach with demonstration video

The English version of the online sharing White Book (Google Doc) can be accessed via the link: <https://docs.google.com/document/d/1EJwwQGUPNtAS1BvjsMvYE423xOnFWwmd_F4UEhfnKkk/edit?usp=sharing>.

The original Chinese version of the document (Tencent Sharing Document) can be accessed via the link: <https://docs.qq.com/doc/DUXNlRnV4Y2xyQkFh>.
