## Supplementary material for "Mapping total microbial communities and waterborne pathogens in household drinking water in China by citizen science and metabarcoding": Supporting Information.docx

Note: All the supplementary tables are provided in a separate Excel file (Supporting Table.xlsx).

### S1 Table. Sample information.

### S1 Text. Top 20 ASVs in the three temperature categories.

Samples in each temperature category were dominated by at least two of ASV14 (genus NA), ASV18 (genus *Sphingomonas*), ASV10 (genus *Porphyrobacter*), and ASV20 (genus *Blastomonas*). However, the most abundant ASV in each T category belonged to different classes: *Blastocatellia* for High T, *Gammaproteobacteria* for Medium T, and *Planctomycetes* for Low T.

### S1 Fig. Relative abundance of top 20 ASVs in household drinking water samples.

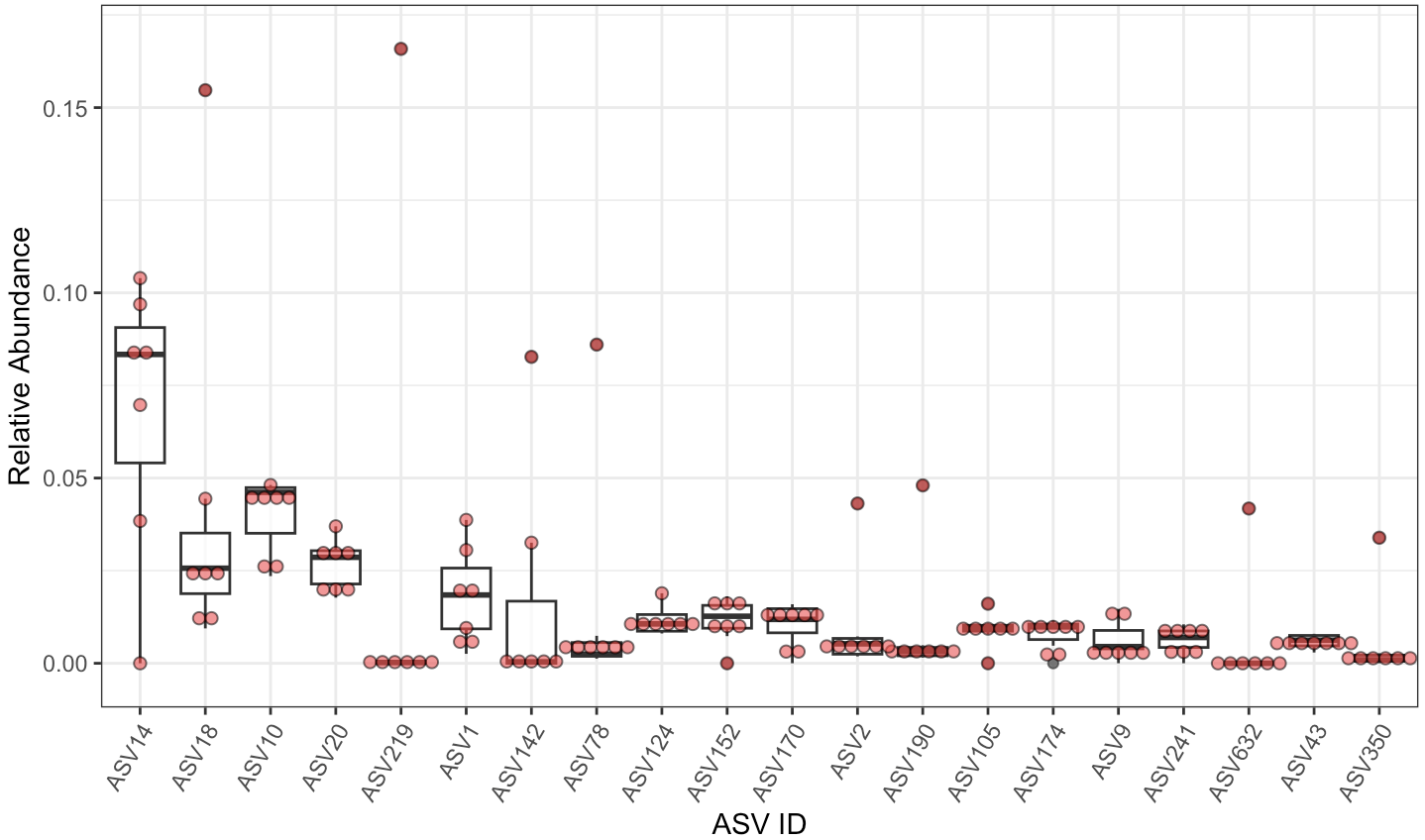

**(A) High temperature**

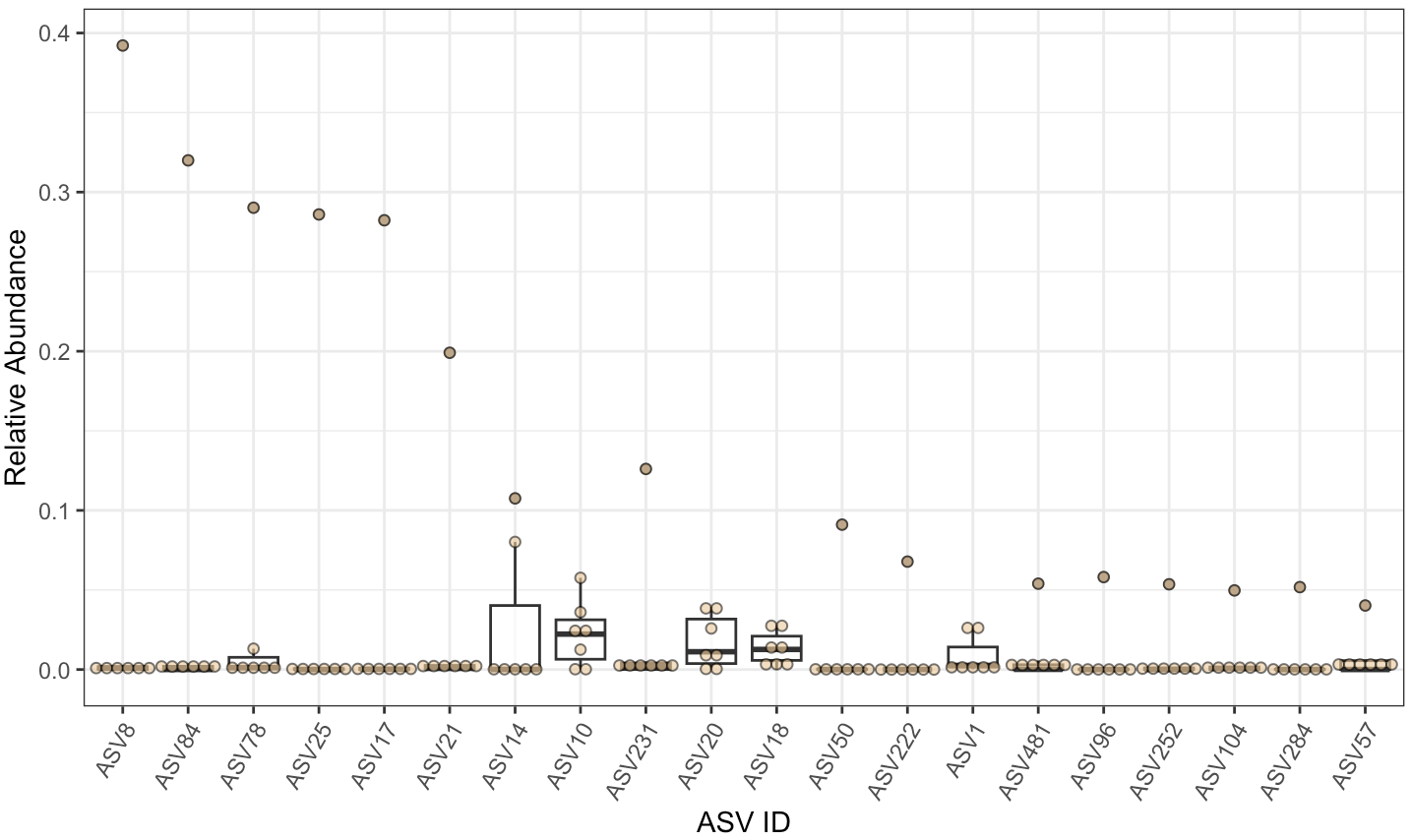

**(B) Medium temperature**

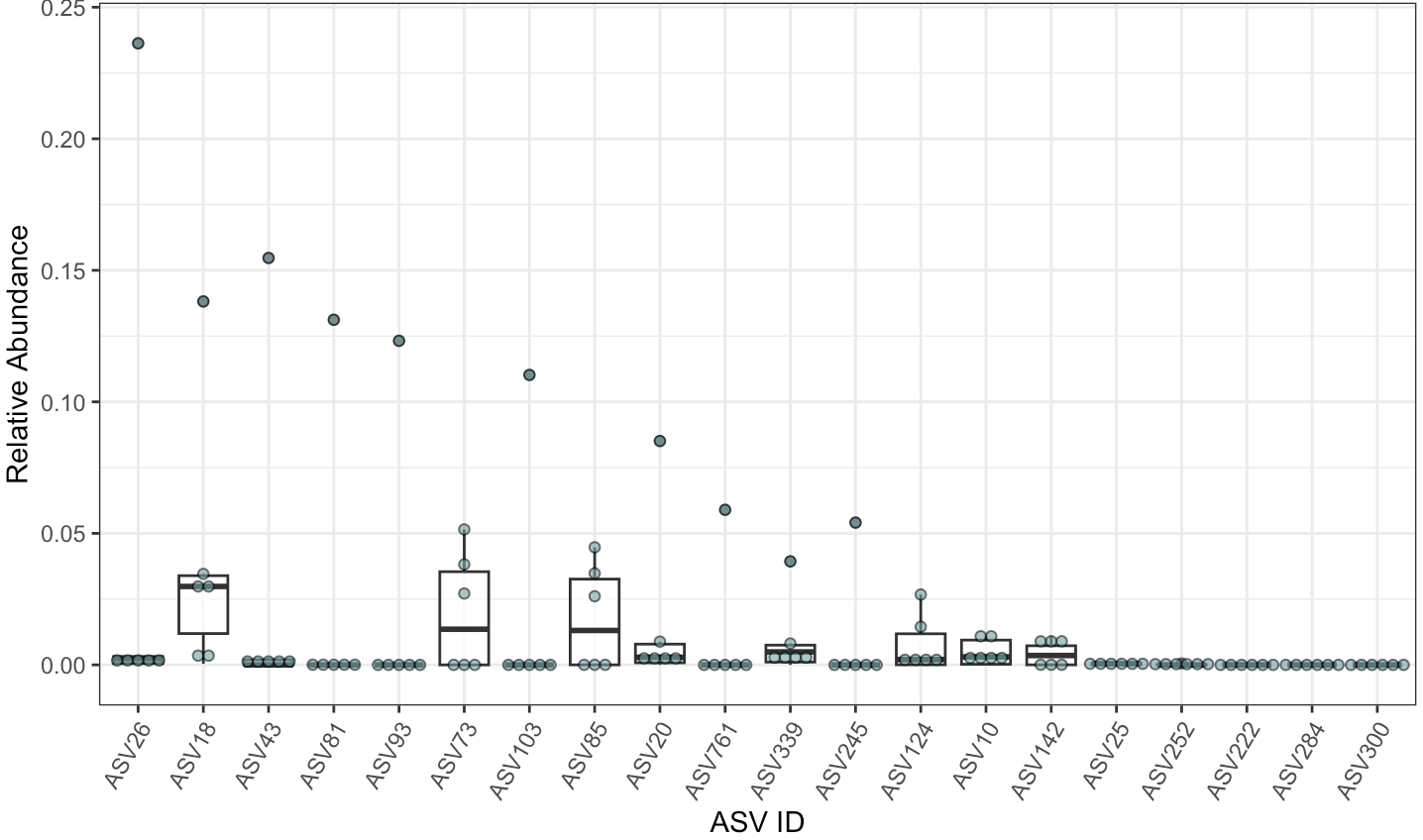

**(C) Low temperature**

### S2 Table. Fisher’s exact test: Positive PCR signal vs. Negative PCR signal in terms of annual precipitation (non-humid and humid).

### S3 Table. The phylum-level taxonomic composition for each sample.

### S2 Text. Beta-diversity (Normal samples, n = 20).

The beta diversity of the total microbial communities in the normal samples are shown in Figure S2. Principal Axis 1, 2, and 3 for PCoA (Bray-Curtis dissimilarity) represent 22.2%, 15.1%, and 8.8% of the variation among the samples respectively. According to PERMANOVA and PERMDISP, the influence of outdoor temperatures was statistically significant (R^2^ = 0.24, adjusted P = 0.015 between High T and Low T), and the differentiation was not due to differences in group dispersions (P = 0.132) (S4 Table). Interestingly, some samples from different regions were highly similar in microbial community composition and structure. For example, SH_0411 & XA_0217, CZ_0712 & TJ_0715.

### S2 Fig. Principal coordinates analysis (PCoA) of the normal samples (n = 20).

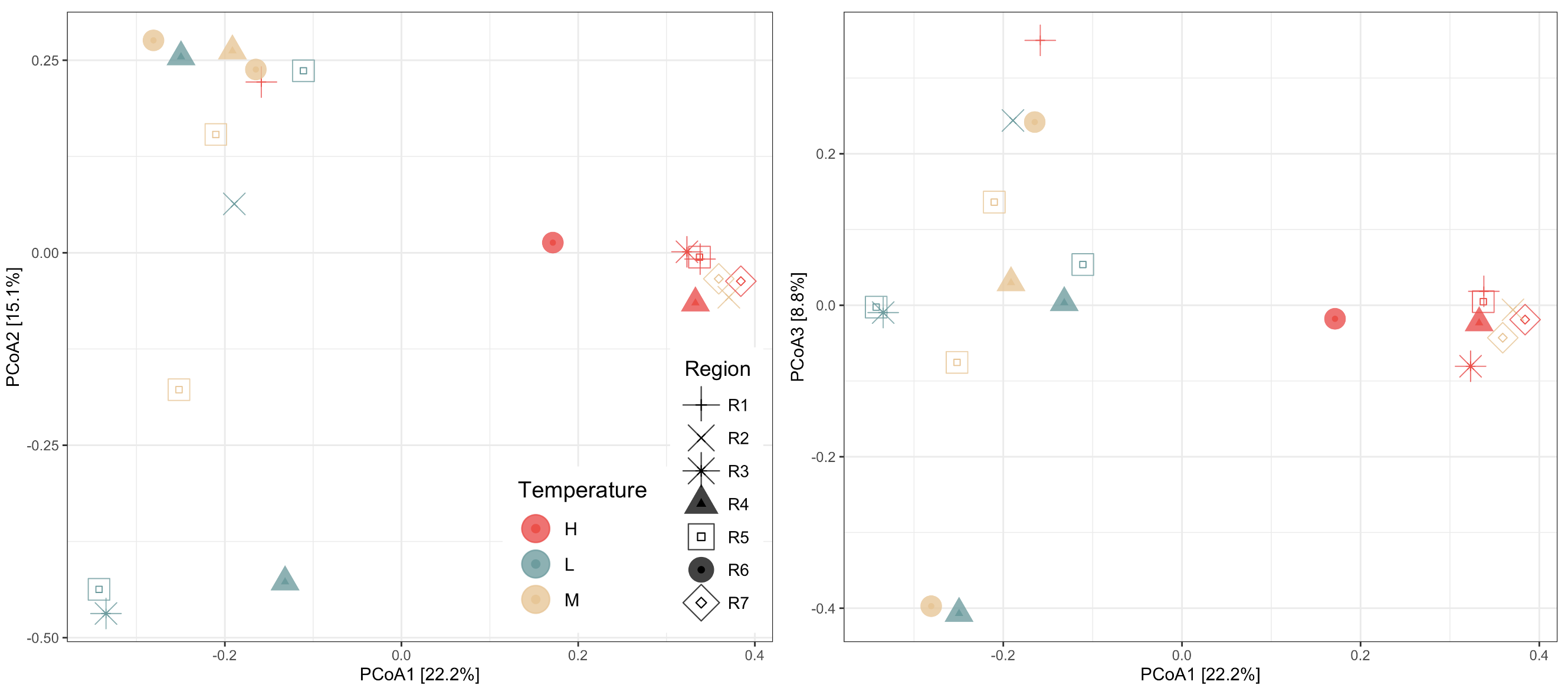

Based on the distance matrix of Bray-Curtis dissimilarity of microbial profiles (ASV-based) among the normal samples. PERMANOVA and post-hoc statistical test results are provided in S4 Table.

### S4 Table. PERMANOVA and PERMDISP of beta diversity.

### S5 Table. Mantel statistic (Spearman's rank correlation, permutations = 999).

### S6 Table. The relative abundance of bacterial pathogens in each sample.

### S7 Table. BLAST+ results of potential pathogens.

### S8 Table. Pathogens in drinking water systems and their related diseases.

### S3 Text. Sample storage test.

#### S3.1 Procedure

Owing to the unanticipated inundation in Zhengzhou, the delivery service within Henan Province experienced a suspension. Consequently, the volunteers were precluded from conducting immediate filtration of the collected water samples without the sampling kit. Despite this impediment, volunteers were instructed to collect duplicate tap water samples in unused mineral water bottles, one to be stored at 4°C and the other at -20°C, pending the arrival of the sampling kit. Subsequently, the water samples were preserved for a duration of 10 days prior to filtration. It is noteworthy that scant scholarly inquiry has addressed the circumstance wherein water samples undergo filtration subsequent to an extended period of storage. Therefore, in order to discern the ramifications of such sample storage under varying temperatures and durations on microbial concentrations within water samples, a series of sample storage tests were conducted within the laboratory.

An initial assessment was conducted utilizing tap water samples obtained from the laboratory tap and surface water samples procured from an artificial water feature at DKU. While certain surface water samples exhibited a positive polymerase chain reaction (PCR) signal, repeated trials yielded no positive PCR signals for any tap water samples, thereby indicating the absence of detectable environmental DNA in DKU tap water. Consequently, to simulate potential contamination in tap water, samples were prepared through the dilution of tap water with surface water from DKU, and these were employed for the subsequent sample storage tests.

During the storage test, the mixed water samples were divided into three groups, each comprising two samples, and were stored in the refrigerator for intervals of 3 days and 7 days, respectively. In each group, one sample was stored at 4°C, and the other at -20°C. After storage, sample filtration, DNA extraction, and PCR amplification were conducted to ascertain the impact of storage conditions on microbial detection.

#### S3.2 Storage Condition Influence

A discernible pattern emerged in the test results: when no bacteria were initially detectable in the tested tap water samples, storage at -18°C effectively prevented bacterial growth up to 7 days of storage. In contrast, samples stored at 4°C yielded positive signal after 3 days. This observation implied that samples stored at 4°C could experience active bacterial growth, and therefore are not suitable for preserving the water samples for microbiome analysis.

*Sample Storage Test PCR Results Summary*

|  | **Immediate Extraction** | **3-Day Storage** | **7-Day Storage** |
| --- | --- | --- | --- |
| **4°C** | Negative | Positive | Positive |
| **-20°C** | Negative | Negative | Negative |

### S4 Text. Representative pathogen species.

#### S4.1 *Mycobacterium* spp.

The atypical *Mycobacterium* spp. can be naturally found in various water environments and are resistant to disinfection (WHO, 2017). As a result, drinking-water supplies are a plausible source of infection. According to Tang et al. (Tang et al., 2021), mycobacteria gene markers were still detected in drinking water that has been treated with 1.5 - 2.3 times the usual dose during hot weather and extreme rainfall period in Jiangsu. In addition, mycobacteria are not detected by HPC techniques and therefore their presence/absence cannot be indicated by the commonly used indicator organism for microbiological quality of drinking water such as *E. coli* (WHO, 2017)*.*

In our study, pathogenic NTM was widely detected in sampled tap water, especially in the samples collected from Nanjing, Changzhou, Shanghai, and Zibo. The increasing incidence and prevalence of infections and deaths from NTM diseases have risen globally (Ratnatunga et al., 2020). Notably, *M. avium* is on the Fifth Contaminant Candidate List (CCL5) drafted by the US EPA as well as one of the members of *M. avium* complex (MAC), a group of pathogenic bacteria related to tuberculosis (TB).

Although historical and contemporary evidence shows that the typical mycobacteria only have human or animal reservoirs, a tiny amount of *M. tuberculosis* which is the etiological agent of TB was detected in the two weather samples. This finding can be supported by a review article which concluded that *M. tuberculosis* is relatively resilient to environmental stresses and could survive for a long time in water and soil (Martinez et al., 2019).

#### S4.2 *Acinetobacter* spp.

*Acinetobacter* spp. are usually present in treated drinking-water supplies but are sensitive to disinfection. They are opportunistic pathogens and there is no confirmed association between their presence in drinking water and clinical diseases such as GI infection among the general population. However, they might cause pneumonia and wound infections in susceptible patients as well as neonates and elderly individuals. Like mycobacteria, *E. coli* or thermotolerant coliforms are not suitable indicators for their presence/absence (WHO, 2017). Small amount of the pathogenic species *A. baumannii* was only detected in the two samples collected after the extreme rainfall events, demonstrating its occasional occurrence and the potential impact of the typhoon and flood. It is part of the multidrug-resistant *A. calcoaceticus baumannii* complex (ACB complex) that causes nosocomial infections (WHO, 2017).

#### S4.3 *Legionella* spp.

*Legionella* spp. are ubiquitous inhabitants in water environments. They generally occur in low numbers in freshwater environments but thrive in some human-made water environments such as cooling towers, hot water distribution systems, and spas, especially in buildings with large, complex plumbing systems. *Legionella* spp. are sensitive to disinfection of monochloramine in particular and not detected by HPC techniques (WHO, 2017).

Almost all species in the genera *Legionella* are thought to be potential human pathogens, but *L. pneumophila* (on CCL5) is the main cause of Legionnaires' disease (pneumonia) and Pontiac fever (a milder infection)*.* Commonly, people get infections when they breathe in small droplets that contain pathogenic *Legionella*. Here, *L. pneumophila* was likely to be detected in 22.7% (5) of the tap water samples, including the ones from Zhengzhou, Macau, Shanghai, Nanchang, and Xi’an. Moreover, some other pathogenic species such as *L. oakridgensis* and *L. maceachernii* occurred in the samples from Tianjin, Lanzhou, Xi’an, Lijiang, and Zhengzhou.

Since *Legionella* spp. can proliferate at temperatures between 25 and 50 °C, water temperature is a crucial factor in control strategies. In addition, attention should also be taken to keeping plumbing systems clean and flowing as well as selecting plumbing materials that are less likely to support excess growth of *Legionella* spp. and the development of biofilms (WHO, 2017).

#### S4.4 *Eschericha* coli

*Eschericha coli* can normally be found in abundance in humans’ and animals’ intestinal flora where it is harmless in general. However, a few enteropathogenic strains account for diarrhea ranging from mild to highly bloody with considerably high infectivity while children under five are at highest risk. In addition, *E. coli* can cause serious diseases in other parts of the body such as meningitis and bacteremia (WHO, 2017). *E. coli* is the common fecal indicator bacteria and the traditional tests can be generally reliable; however, potential viable but non-culturable *E. coli* cells can result in underestimations of the actual water contamination (Liu et al., 2008).

Drinking-water supplies can be contaminated by rainwater runoff that contains cattle excreta (WHO, 2017). The most polluted tap water samples in our study are from Changzhou, Beijing, and Zhengzhou. The elevated RA of *E. coli* in the sample collected after the typhoon event suggests that the pathogen contamination might be related to typhoon.

#### S4.5 *Salmonella* spp.

*Salmonella* spp. are frequently present in surface and drinking water (Liu et al., 2018). However, they do not originate in water, and thus their presence denotes fecal contamination (Annous and Gurtler, 2012). *Salmonella* spp. are usually transmitted by the ingestion of contaminated water or food (Knodler and Elfenbein, 2019), causing a variety of diseases ranging from acute gastroenteritis to fatal typhoidal fever (Dekker and Frank, 2015). In particular, the waterborne typhoid fever outbreak is a very serious global public health problem.

*Salmonella* spp. are relatively sensitive to disinfection; however, a tiny proportion of *Salmonella enterica* occurred in the post-flood tap water sample from Zhengzhou. Whether this would pose serious health threats to the residents remains uncertain, and the severity of the disease depends on the serotype and host factors of *Salmonella* which were unavailable. However, the presence of *Salmonella enterica* indicates potential fecal contamination in the household drinking water after 2021 Henan Floods.

### S5 Text. Potential Environmental Drivers: Extreme Precipitation and Heat

The detection of a wide range of pathogens in post-weather tap water samples indicates the impacts of extreme rainfall events on urban water safety and public health, while the recent devastating floods in Greece and Hong Kong – Shenzhen further underscore the escalating frequency and intensity of extreme rainfall events globally (Bettiza and Ertl, 2023, Ng, 2023). Stormwater runoff can directly pollute drinking water sources and excessive urban wastewater strains treatment plants, causing untreated sewage discharge into waterways (Langeveld et al., 2013). Eventually, this could lead to drinking water system contamination, which has been supported by the analysis of tap water microbiome and pathogen changes in this research.

S6 Table displays the impact of typhoons and/or floods on tap water pathogen profiles, notably increasing specific species' RA and total species count. Post-weather samples from Changzhou and Zhengzhou showed elevated potential pathogens, ranking second (CZ_0728, 20.7%) and third (ZZ_0802, 10.7%) in RA (Fig 6). Furthermore, both cities displayed increased pathogen species after extreme weather, with some pathogen species (*Salmonella enterica* and *Aeromonas hydrophila*) detected exclusively in these post-event samples. Therefore, the rising risk of waterborne pathogens in post-flush tap water samples indicates that extreme rainfall events are a prominent risk factor in public health (Phan and Sherchan, 2020). To mitigate these risks, investing in wastewater treatment plant upgrades to handle increased effluent and fortifying drinking water treatment and distribution systems against extreme weather-induced pollution is crucial, necessitating emergency response planning and maintenance for resilient systems (Phan and Sherchan, 2020).

Temperature also significantly impacts waterborne pathogen proliferation in freshwater sources. IPCC reports (IPCC, 2008) link rising surface water temperatures to global atmospheric warming since the 1960s across Asia, North America, and Europe. Studies by Lipp et al. reveal that higher temperatures create favorable conditions for bacteria less sensitive to temperature fluctuations (Lipp et al., 2002), promoting the growth of pathogenic species like *cholera* (*V. cholerae*) and various *Vibrio* species (Funari et al., 2012). Our study supports these connections, demonstrating that warmer weather potentially drives pathogen profiles in tap water. Specifically, we observed a positive correlation between outdoor temperatures equal to or higher than 17°C and RA of potential pathogens, though statistically insignificant given the small sample size (n = 12, R = 0.46, P = 0.13) (S15 Fig). Notably in Changzhou, the summer sample CZ_0712 (outdoor T: 33.3°C) exhibited much higher pathogen abundance and species count (20.7%, 7 genera) compared to the winter sample CZ_0220 (1.80%, 4 genera, outdoor T: 17.0°C).

Extreme rainfall and high temperatures, interconnected, can create a compounding effect that worsens waterborne disease spread. The IPCC AR6 report forecasts continual temperature rise, surpassing 1.5°C and 2°C global warming thresholds without significant emissions reductions. It emphasizes that each temperature increase amplifies extreme weather impact, intensifying heatwaves and heavy rainfall (Masson-Delmotte et al., 2021, Team et al., 2023). Securing tap water safety will become increasingly demanding amid climate change's escalating effects, necessitating stringent control and maintenance.

### S3 Fig. Linear correlation (normal samples, Spearman): Outdoor temperature (higher than 17°C) ~ Total relative abundance of potential pathogens.

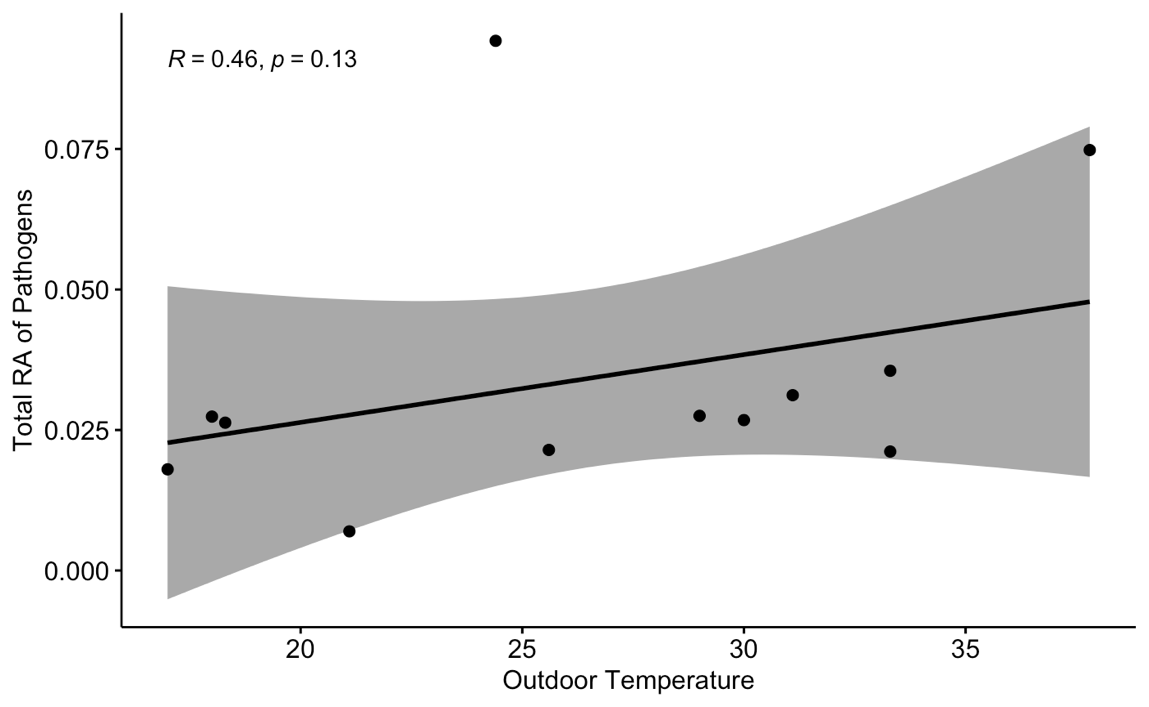

WHO 2017. Guidelines for Drinking-Water Quality (Fourth Edition).
