## Supplementary figures and images for "Mapping total microbial communities and waterborne pathogens in household drinking water in China by citizen science and metabarcoding"

### S1A Fig.jpg

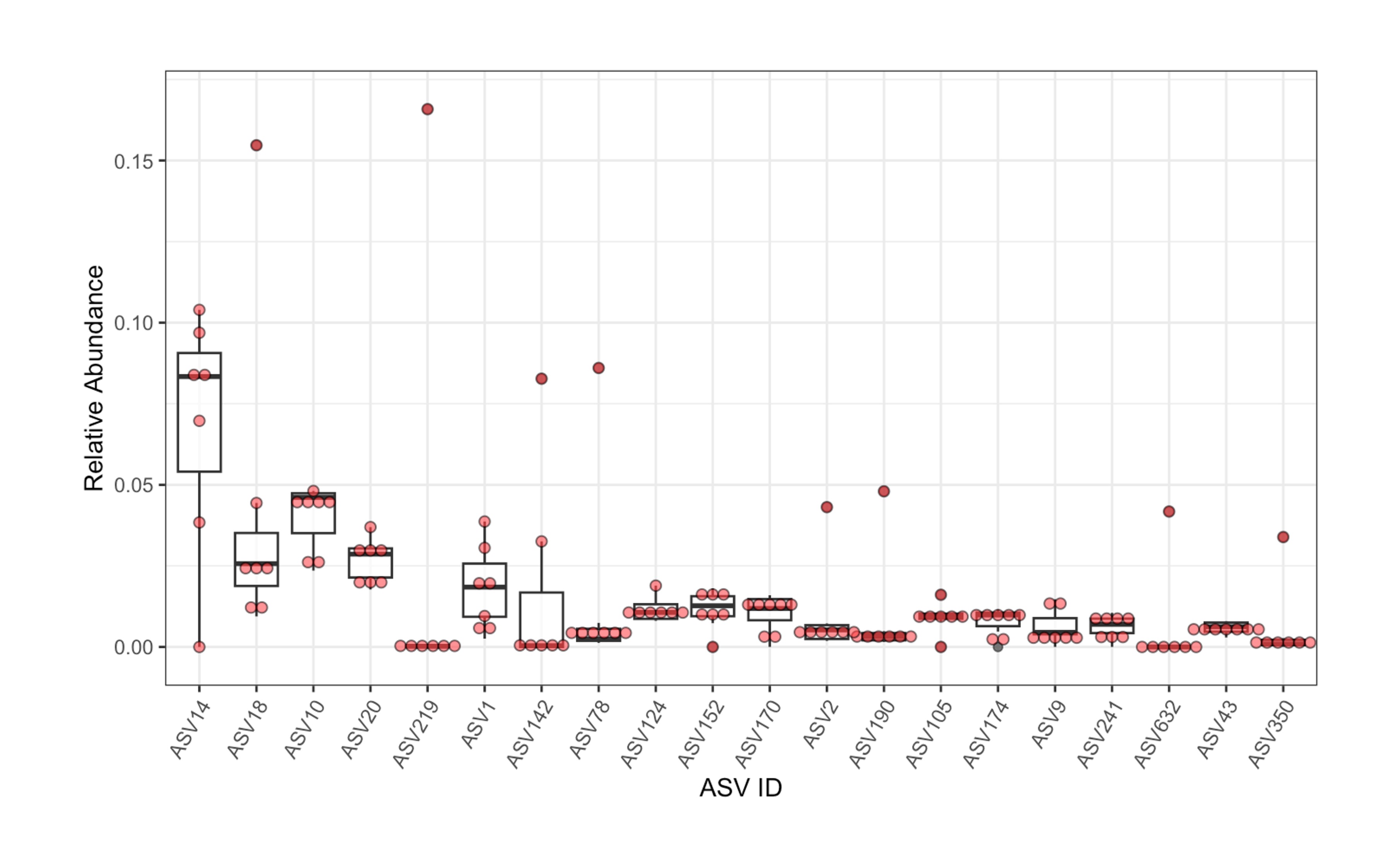

### S1B Fig.jpg

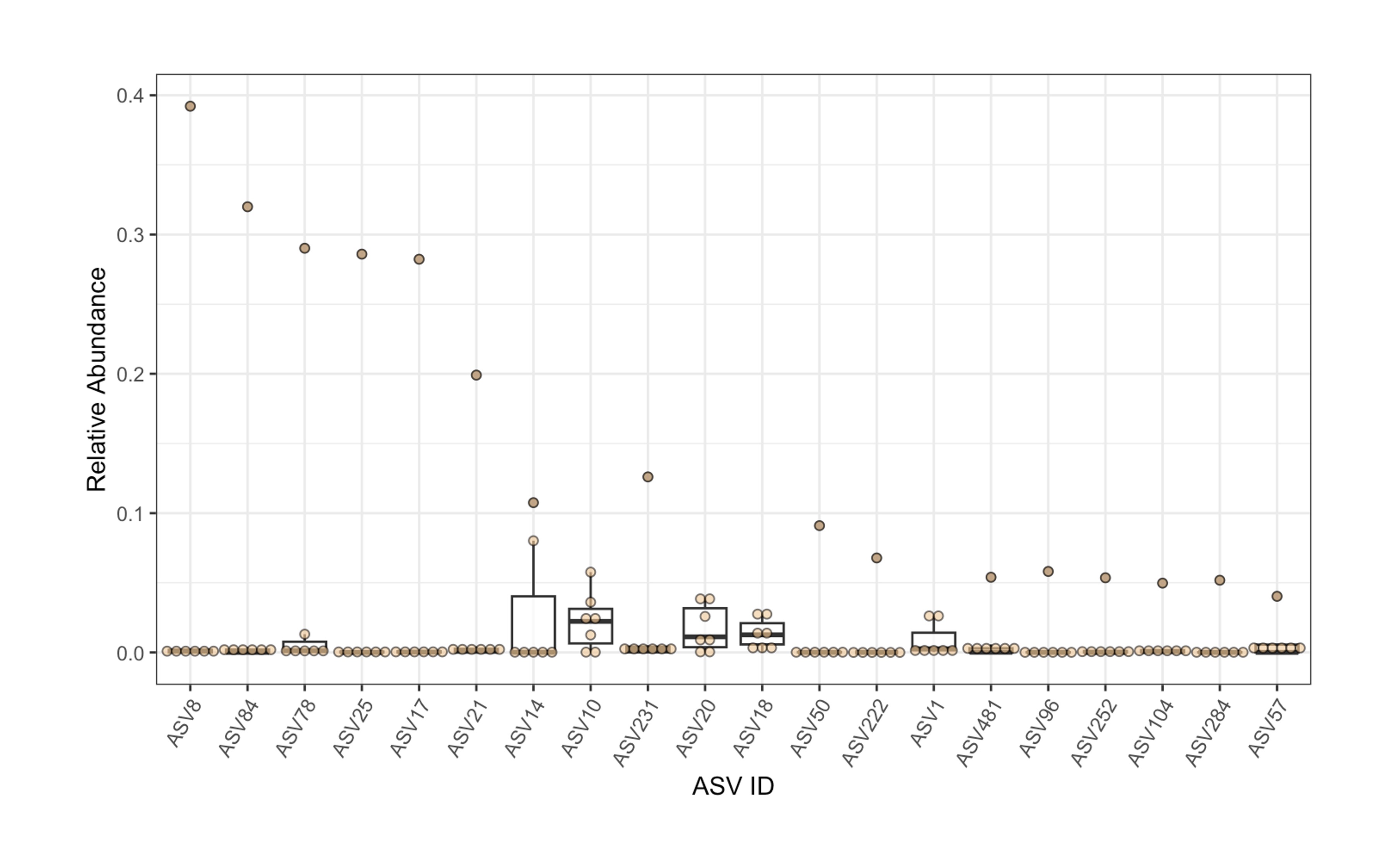

### S1C Fig.jpg

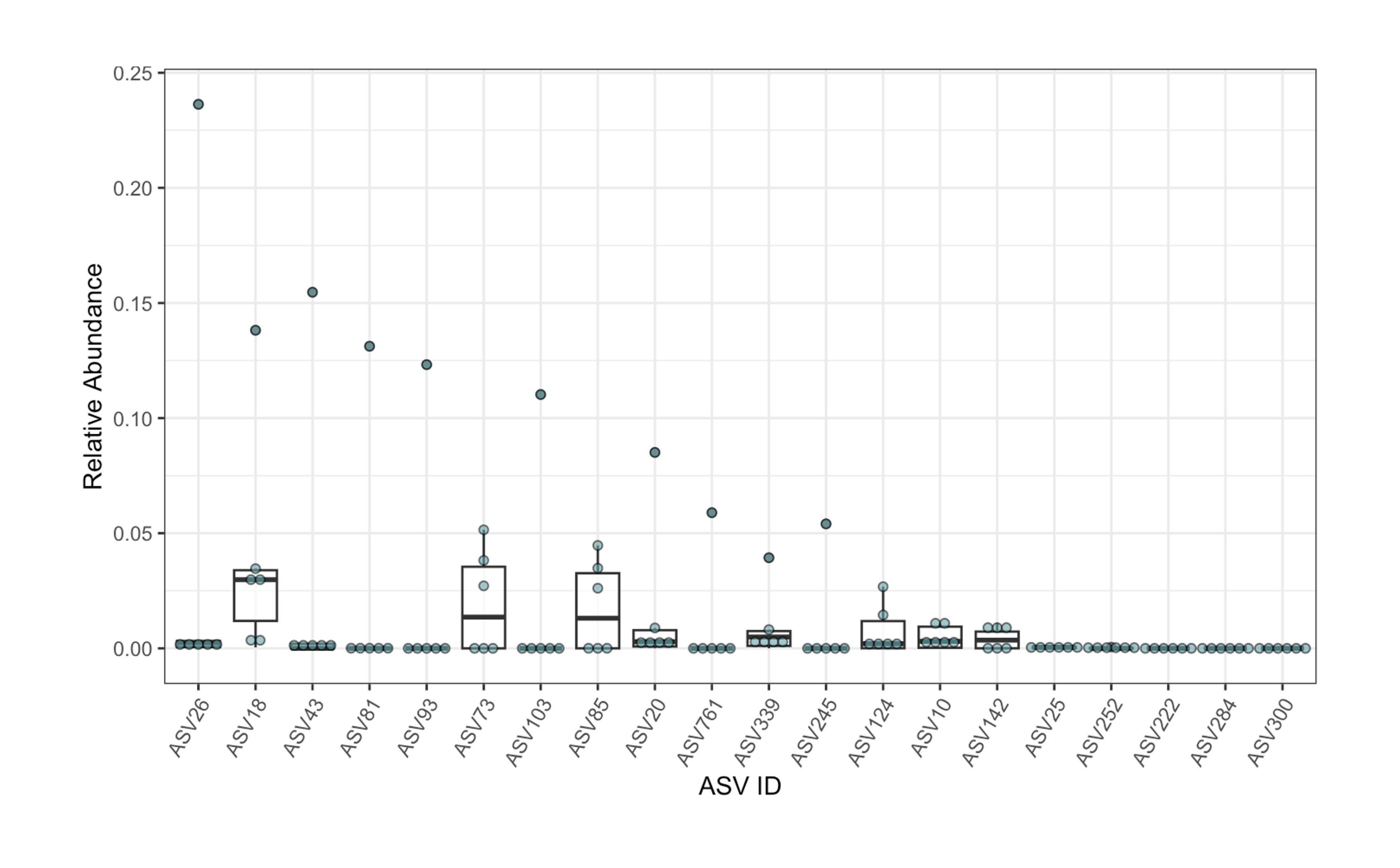

### S2 Fig.jpg

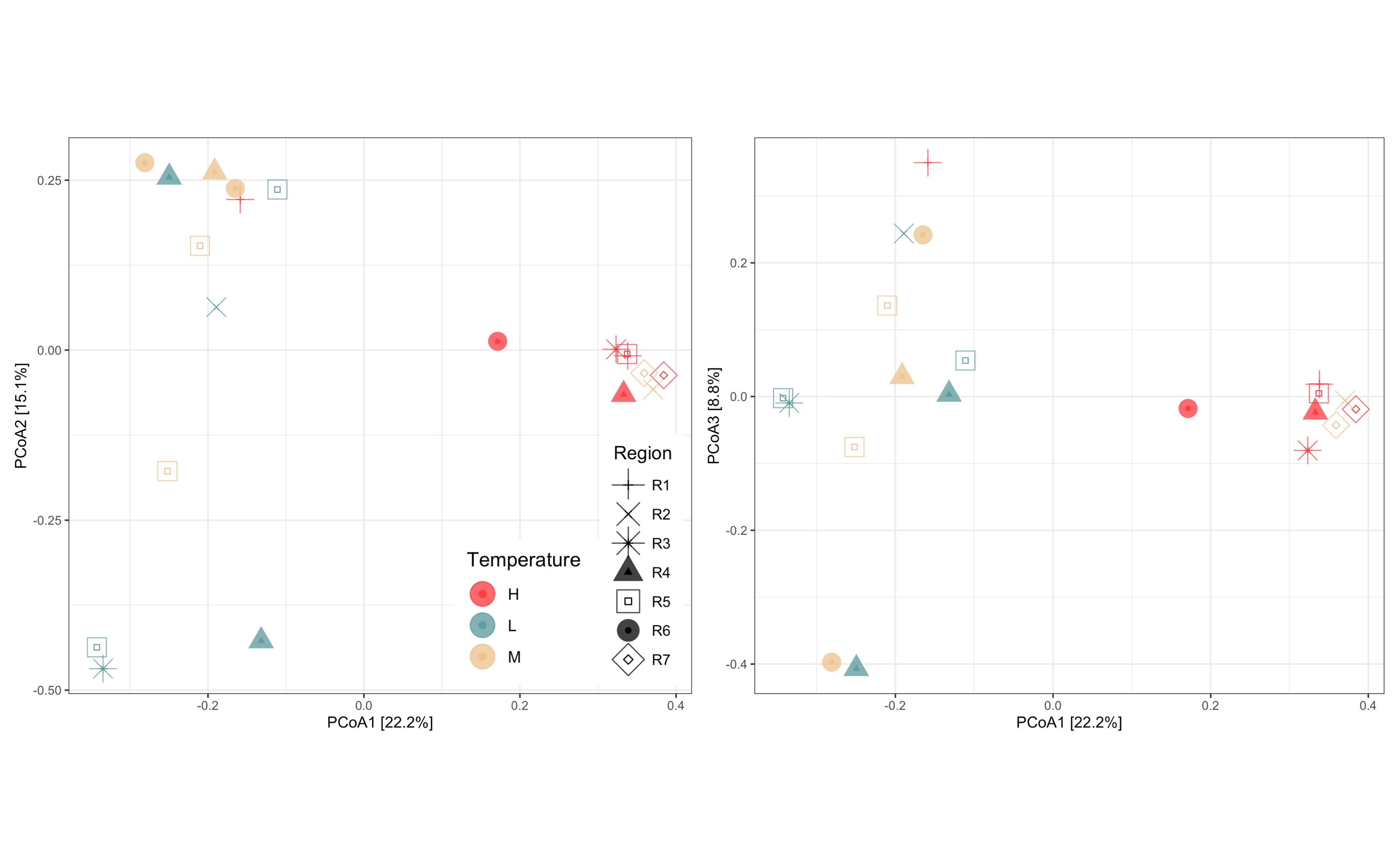

### S3 Fig.jpg

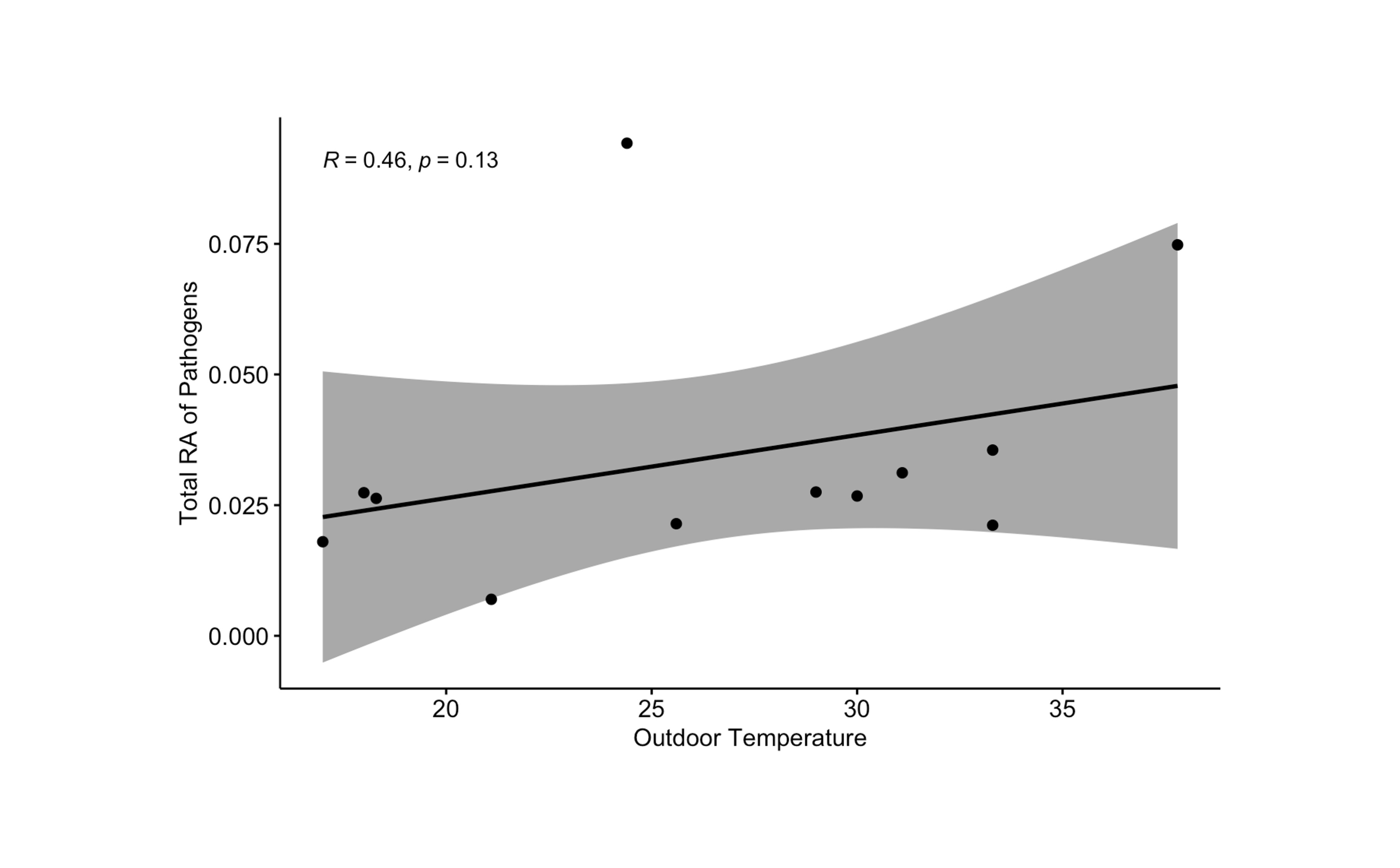
